# Abstraction of Longitudinal Breast Oncology Records Using a Retrieval Pipeline with General-Purpose Large Language Models

**DOI:** 10.64898/2026.03.23.26349012

**Authors:** James C. Dickerson, Marni B. McClure, Margaret Shaw, Marissa B. Reitsma, Nicole H. Dalal, Christina Curtis, Allison W. Kurian, Jennifer L. Caswell-Jin

## Abstract

Manual chart abstraction is a major bottleneck in clinical research. In oncology, important outcomes are often documented only in clinical notes. We developed an open-source pipeline that, in a HIPAA-compliant setting, uses general-purpose large language models (LLMs) to abstract variables from longitudinal patient records without fine-tuning or hand-selecting input documents. We randomly selected 100 patients from a complex institutional breast cancer cohort and compared four LLMs with oncologist abstraction, genetic testing records, and, for select variables, research coordinator abstraction. The unedited charts contained a median of 3,100 pages of text, 7 lines of therapy, and 6.5 years of follow-up. Across the two best-performing models, GPT-5 and Gemini 2.5 Pro, the pipeline achieved 90%–100% concordance for recurrence, BRCA1/2, hormone receptor, HER2, clinical stage, PIK3CA, and ESR1 status. Anti-cancer drug identification was comparable to that of a second oncologist. Therapy-line reconstruction was less accurate than a second oncologist; all four LLMs outperformed research coordinators on both tasks. In a separate cohort of 97 patients, the unmodified pipeline showed similar performance for recurrence detection. These findings suggest that LLMs may enable scalable abstraction from narrative medical records, but replication and validation across institutions and diseases are needed.

## Introduction

Clinical research depends on abstracting key variables from the electronic medical record and converting them into structured data for analysis.^1,2^ This manual process is how we build disease registries, retrospective cohorts, and many real-world evidence datasets. Abstracting information from the chart is labor-intensive and vulnerable to inconsistency across reviewers. Many key variables such as diagnoses, outcomes, biomarker results, treatment exposures, and disease progression are often documented only in free text rather than in structured fields. As a result, much of the clinically meaningful information in the narrative record is not currently used for research, which limits the completeness and quality of observational datasets.^3,4^

Breast cancer research illustrates this problem. Most metastatic breast cancer cases are recurrences of early-stage disease, but SEER only captures initial treatment for patients with *de novo* metastatic disease, who represent just 6% of new diagnoses.^5,6^ As a result, population-level estimates of metastatic breast cancer burden in the United States depend on simulation models and claims-based inference.^5,7,8^ Retrospective studies that inform clinical decision-making often rely on labor-intensive manual abstraction of smaller-than-ideal cohorts because identifying cancer recurrence is challenging at scale.^9,10^ Answering the full range of clinically meaningful questions through manual chart review is impossible.

Large language models (LLMs) may offer a practical way to automate this work. To date, approaches to clinical data extraction have included both task-specific models trained on institutional and public data and commercially available general-purpose LLMs (e.g., ChatGPT, Gemini) used to extract data from human-selected documents.^11–14^ We therefore sought to understand whether a system that autonomously retrieved relevant documents from the full longitudinal record, coupled to general-purpose LLMs in a HIPAA-compliant environment, could reliably abstract a broad range of variables from complex longitudinal cancer records. The goal was to minimize both the per-patient human level work and the technical work needed to run the system: no manual selection or annotation of inputs, fine-tuning, labeled training data, institution-specific retraining, or large on-premises computational resources.^15^ If successful, this approach could provide a practical path toward unlocking larger volumes of clinical data for research.

We developed a chart abstraction pipeline using commercially available LLMs and applied it to patients with complex breast cancer histories (Figure 1). We compared LLM-derived abstractions across multiple models with oncologist review for recurrence, systemic therapy, tumor characteristics, and genomic testing. Given the complexity and potential subjectivity of systemic therapy abstraction, we benchmarked this task against a second oncologist and research coordinators. We then assessed whether LLM-derived datasets preserved downstream survival estimates. Lastly, we used the unmodified pipeline and prompts to detect recurrence events and adjuvant endocrine therapy use in an independent cohort of 97 patients who were diagnosed with early-stage breast cancer and then became pregnant.

**Figure 1.**
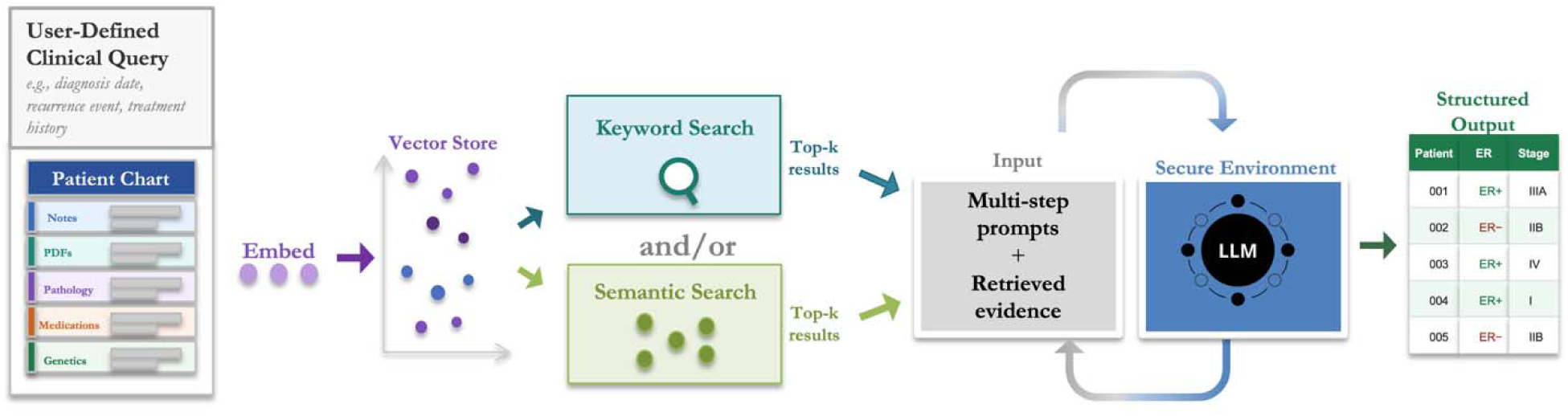
Fully Automated Chart Abstraction Workflow. Text documents from the electronic medical record (Epic) were exported from the institutional research warehouse, segmented into chunks, and indexed for exact-word, BM25, and semantic retrieval. For each task, retrieved text was then passed to commercially available LLMs using schema-constrained prompts to generate structured outputs.

## Results

### Overview and cohort characteristics

For simplicity, the main text focuses on the two LLMs that performed best overall (Gemini 2.5 Pro and GPT-5); results for all four models are in Supplementary Figure 1 and Supplementary Table 1.

The 100-patient cohort was complex with around 1,100 individual drugs prescribed in a median of 7 lines of therapy (interquartile range [IQR], 4.75 – 9) over 6.5 years of follow-up from diagnosis (IQR, 3.7 – 9.9 years). The median chart size was approximately 2.3 million tokens or roughly 3,100 pages of text (Table 1). The average time to process an individual patient ranged from 1 – 11 minutes (Supplementary Table 2).

**Table 1.** Cohort characteristics. Clinical characteristics from the oncologist derived dataset of the 100-patient breast cancer cohort and measures of chart complexity and size.

| Characteristic | Value |
| --- | --- |
| **Demographic and disease characteristics (n=100)** |  |
| Age at diagnosis, median (IQR), y | 46.5 (40.3 – 56.9) |
| Stage at diagnosis, No. (%) |  |
| 1 | 15 (15%) |
| 2 | 37 (37%) |
| 3 | 32 (32%) |
| 4 | 16 (16%) |
| Tumor subtype at diagnosis, No. (%) |  |
| HR+/HER2– | 55 (55%) |
| HR+/HER2+ | 8 (8%) |
| HR–/HER2+ | 5 (5%) |
| Triple-negative | 32 (32%) |
| Germline BRCA1/2 pathogenic variant present, No. (%) | 3 (3%) |
| Somatic PIK3CA mutation present, No. (%) | 30 (30%) |
| Somatic ESR1 mutation present, No. (%) | 18 (18%) |
| **Recurrence Events (n=84; excludes stage 4 patients)** |  |
| Any recurrence (locoregional or distant), No. (%) | 82 (98%) |
| Locoregional recurrence, No. (%) | 28 (33%) |
| Distant metastatic recurrence, No. (%) | 78 (93%) |
| **Survival status** |  |
| Documented death by censor date, No. (%) | 49 (49%) |
| Documented follow-up from diagnosis, median (IQR), y | 6.5 (3.7 – 9.9) |
| **Systemic therapy complexity** |  |
| Total lines of systemic therapy abstracted, No. | 716 |
| Lines of systemic therapy per patient, median (IQR) | 7 (4.75 – 9) |
| **Chart complexity** |  |
| Estimated chart size, median (IQR), MB | 8.9 (4.6 – 15.5) |
| Estimated tokens per chart, median (IQR) | 2.3 (1.2 – 4.1) million |
| Estimated pages per chart, median (IQR) | 3,100 (1,624 – 5,419) |
| Clinical notes per patient, median (IQR), No. | 426 (218 – 695) |
| Pathology documents per patient, median (IQR), No. | 6 (4 – 11) |
| Distinct primary treating oncologists across cohort, No. | 20 |

### Agreement for non-systemic therapy variables

Across non-systemic variables, the pipeline showed high agreement with expert oncologist abstraction (Figure 2; Supplementary Figure 1; Supplementary Table 1). The two best overall performing LLMs achieved at least 90% agreement across all 13 variables. Agreement was highest for variables anchored to pathology reports or demographic data. Variables with heterogeneous documentation, lower performing retrievers, those requiring longitudinal reasoning such as clinical stage assignment, and variables often available only in scanned PDFs or screenshots not retained in the research warehouse had lower agreement (Figure 2; Supplementary Table 3; Supplementary Table 4).

**Figure 2.**
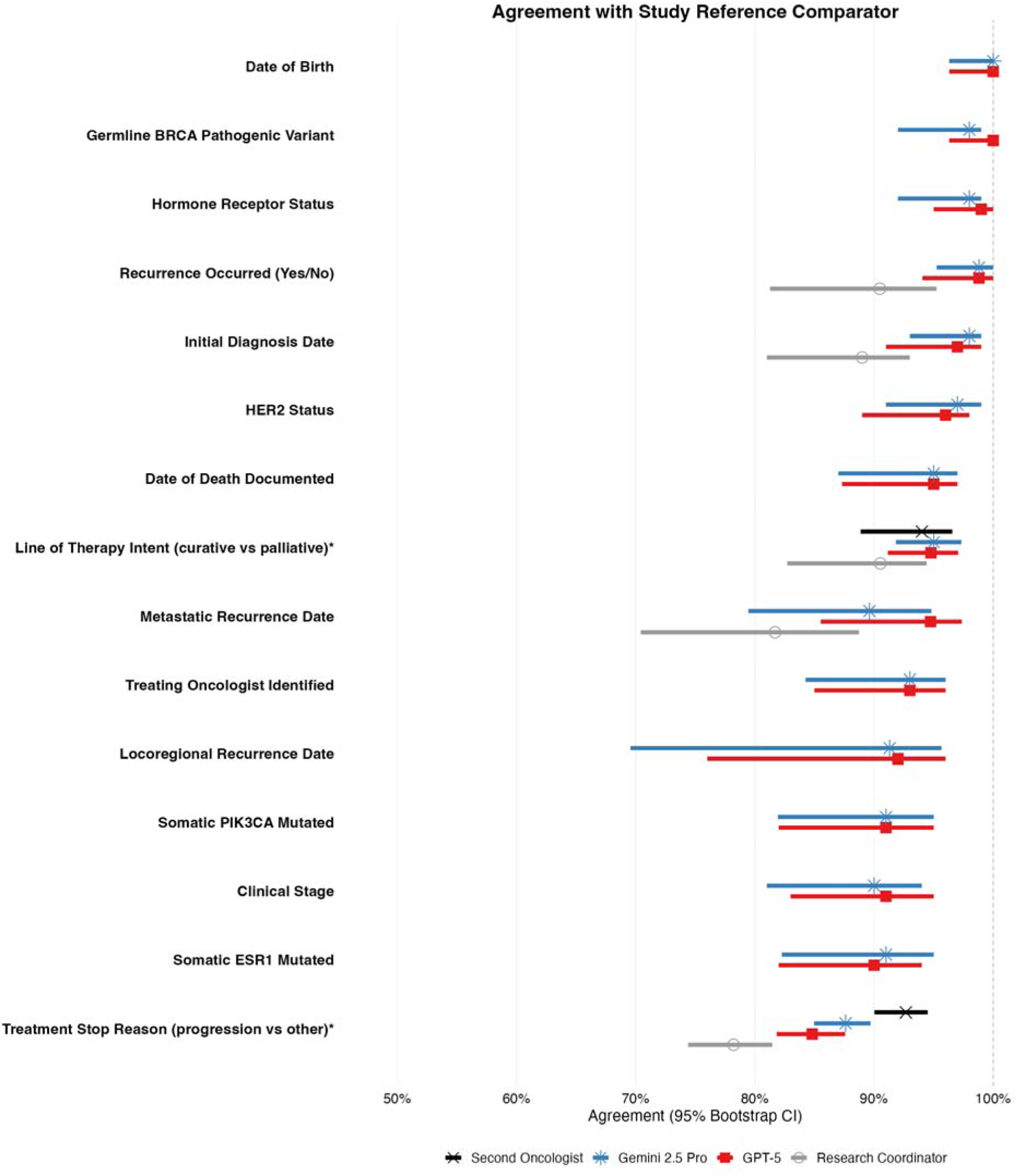
Concordance with oncologist abstraction and linked genetic testing records across key clinical variables. Forest plot showing agreement with expert oncologist abstraction and 95% bootstrap confidence intervals for the two best-performing LLMs (Gemini 2.5 Pro and GPT-5) across key clinical variables. Research coordinators are shown for variables they previously abstracted. Date-based variables were evaluated using concordance within ± 90 days. Categorical variables were evaluated using exact concordance. Reference comparators were linked genetic testing records for germline and PIK3CA mutations and the oncologist for all other variables. The * denotes that treatment intent and reason for discontinuation were evaluated only among matched therapy lines

For the original diagnosis date, the two best-performing LLMs agreed with the expert on 98% and 97% of patients. No original diagnosis dates differed by more than 1 year (Supplementary Figure 2). For metastatic recurrence, agreement within ± 90 days was 90% and 96% for the two best-performing LLMs; 82% and 90% of recurrence dates were within 30 days. Locoregional recurrence had high date concordance (Supplementary Figure 2), but more false negatives than metastatic recurrence. The two best-performing LLMs missed 5 and 3 events, respectively. In the 6 total unique cases, the LLMs had classified the recurrence as distant metastatic rather than locoregional. Full results for the best two models are in Table 2. All four model-by-model results are shown in Supplementary Table 1, with an expanded forest plot in Supplementary Figure 1, confusion matrices for stage and hormone receptor and HER2 status in Supplementary Figure 3, and a qualitative error analysis in Supplementary Table 3. Of 700 patient-variable abstractions based on a single expert comparator, 55 (7.9%) met the criterion for discordance audit and were reviewed by at least two clinicians with consensus adjudication of the final reference label.

**Table 2.** Abstraction performance across evaluated clinical variables. Agreement metrics for the two best-performing overall LLMs, across all variables. CIs were estimated using bias-corrected and accelerated bootstrap resampling at the patient level with 1,000 iterations. Jaccard, recall, precision, and F1 score are presented as per-patient values.

| Clinical domain / variable | Second oncologist | Gemini 2.5 Pro | GPT-5 | Research coordinator |
| --- | --- | --- | --- | --- |
| **Timing of key events** |  |  |  |  |
| Documented death date identified | — | 95%<br>[88 - 97] | 95%<br>[88 - 98] | — |
| Date of diagnosis ($\pm 90$ d) | — | 98%<br>[92 - 99] | 97%<br>[91 - 99] | 97%<br>[90 - 99] |
| Recurrence occurred (yes/no) | — | 99%<br>[93 - 100] | 99%<br>[95 - 100] | — |
| Date of first locoregional recurrence ($\pm 90$ d) | — | 91%<br>[70 - 96] | 92%<br>[68 - 96] | — |
| Date of first metastatic diagnosis ($\pm 90$ d) | — | 90%<br>[81 - 95] | 96%<br>[89 - 98] | 73%<br>[63 - 80] |
| **Clinical information** |  |  |  |  |
| Original HR/HER2 subtype | — | 95%<br>[88 - 98] | 95%<br>[87 - 98] | — |
| Original stage | — | 90%<br>[81 - 94] | 91%<br>[83 - 95] | — |
| Primary treating oncologist | — | 93%<br>[86 - 96] | 93%<br>[86 - 96] | — |
| **Genetic testing** |  |  |  |  |
| Germline BRCA1/2 mutation identified | — | 98%<br>[93 - 99] | 100%<br>[96 - 100] | — |
| Somatic PIK3CA mutation present | — | 91%<br>[83 - 95] | 91%<br>[83 - 95] | — |
| Somatic ESR1 mutation present | — | 91%<br>[82 - 95] | 90%<br>[82 - 94] | — |
| **Systemic therapy** |  |  |  |  |
| All prescribed anti-cancer drugs: Jaccard similarity (mean) | 0.95<br>[0.92 - 0.96] | 0.90<br>[0.87 - 0.92] | 0.91<br>[0.89 - 0.93] | 0.75<br>[0.72 - 0.79] |
| All prescribed anti-cancer drugs: precision | 97%<br>[95 - 98] | 92%<br>[90 - 94] | 93%<br>[91 - 95] | 88%<br>[85 - 90] |
| All prescribed anti-cancer drugs: recall | 98%<br>[96 - 99] | 97%<br>[95 - 98] | 97%<br>[95 - 98] | 83%<br>[79 - 86] |
| All prescribed anti-cancer drugs: F1 score | 97%<br>[96 - 98] | 94%<br>[92 - 95] | 95%<br>[93 - 96] | 85%<br>[82 - 87] |
| Systemic therapy lines: regimen Jaccard (mean) | 0.80<br>[0.76 - 0.84] | 0.70<br>[0.66 - 0.75] | 0.70<br>[0.66 - | 0.49<br>[0.44 - 0.54] |

**Table 2. Abstraction performance across evaluated clinical variables.**
| Clinical domain / variable | Second<br>oncologist | Gemini 2.5<br>Pro | GPT-5 | Research<br>coordinator |
| --- | --- | --- | --- | --- |
|  |  |  | 0.75] |  |
| Systemic therapy lines: exact-content<br>precision (mean) | 88%<br>[84 - 90] | 79%<br>[76 - 83] | 81%<br>[77 - 84] | 62%<br>[57 - 67] |
| Systemic therapy lines: exact-content<br>recall (mean) | 87%<br>[82 - 90] | 82%<br>[78 - 86] | 81%<br>[77 - 84] | 63%<br>[58 - 67] |
| Systemic therapy lines: exact-content F1<br>(mean) | 87%<br>[84 - 90] | 80%<br>[76 - 83] | 80%<br>[76 - 83] | 62%<br>[57 - 66] |
| Start date of matched treatment line<br>($\pm 30$ d) | 98%<br>[96 - 99] | 94%<br>[92 - 96] | 96%<br>[93 - 97] | 95%<br>[91 - 97] |
| Stop date of matched treatment line<br>($\pm 30$ d) | 90%<br>[86 - 92] | 81%<br>[76 - 85] | 79%<br>[73 - 83] | 74%<br>[69 - 78] |
| Treatment intent for matched lines | 93%<br>[89 - 96] | 95%<br>[90 - 97] | 95%<br>[91 - 97] | 91%<br>[86 - 94] |
| Reason for discontinuation for matched<br>lines | 92%<br>[90 - 95] | 87%<br>[83 - 89] | 83%<br>[80 - 87] | 78%<br>[74 - 82] |

### Systemic therapy abstraction relative to inter-expert variability

Compared with the first expert oncologist, the second oncologist achieved a mean patient-level Jaccard similarity of 0.95 (95% confidence interval [CI], 0.92 to 0.96) for all prescribed anti-cancer drugs and 0.80 (95% CI, 0.76 to 0.84) for reconstructing the exact lines of therapy (Figure 3; Supplementary Table 1). Among matched lines (n = 668), oncologist agreement was 93% for treatment intent, 92% for reason for discontinuation, 98% for start date within ± 30 days, and 90% for stop date within ± 30 days.

**Figure 3.**
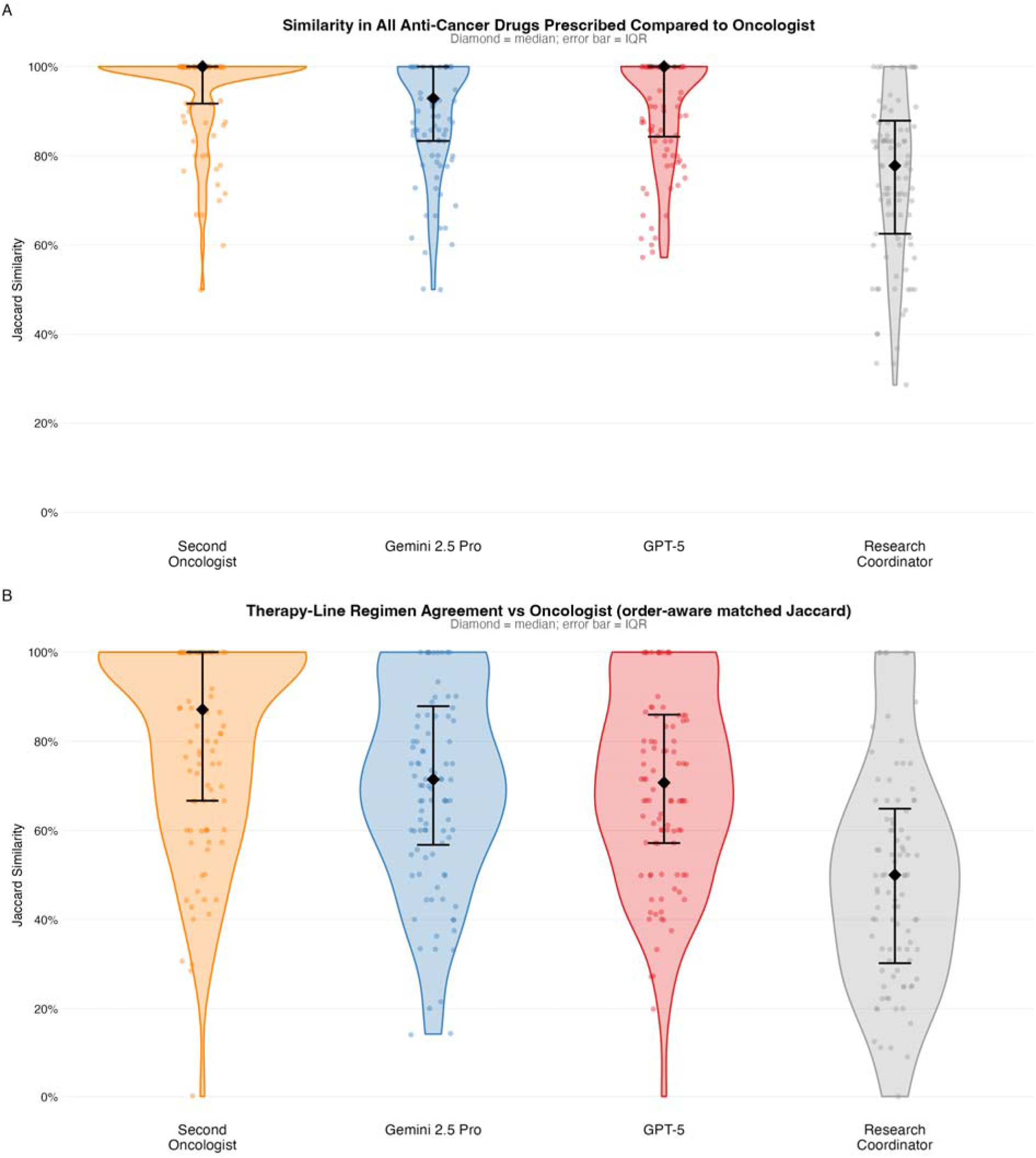
Patient-level overlap in systemic therapy abstraction. Violin plots showing patient-level Jaccard similarity for abstraction of all prescribed anti-cancer drugs (Panel A) and exact therapy lines (Panel B). Diamonds indicate medians and bars are the interquartile ranges. Results are shown for the second oncologist, the two best-performing LLMs, and research coordinators relative to the expert oncologist reference.

The two LLMs had a mean patient-level Jaccard similarity of 0.90 (95% CI, 0.87 to 0.92) and 0.91 (95% CI, 0.89 to 0.93) for prescribed anti-cancer drugs with CIs that overlapped those of the second oncologist. For lines of therapy, they were lower at 0.70 (95% CI, 0.66 to 0.75) and 0.70 (95% CI, 0.66 to 0.75) and the CIs did not overlap. When the two best-performing LLMs were compared with each other, they disagreed to a similar extent as the two expert oncologists did with Jaccard similarities of 0.96 (95% CI, 0.94 to 0.98) for drugs and 0.80 (95% CI, 0.75 to 0.84) for lines of therapy. Violin plots for all four LLMs are shown in Supplementary Figure 4; Supplementary Table 5 shows the most common missed and added drugs.

For exact systemic therapy line reconstruction, precision, recall, and F1 were 79%, 82%, and 80% for Gemini 2.5 Pro and 81%, 81%, and 80% for GPT-5, compared with 88%, 87%, and 87% for the second oncologist (Table 2). Using the start date for line matching (Table 3, Supplementary Table 6) on average the LLMs produced similar amounts of added and subtracted lines of therapy.

**Table 3.** Direction of error in systemic therapy abstraction relative to the expert oncologist reference. The table summarizes line-level error across the second oncologist, research coordinators, and the two best-performing overall LLMs, including correctly matched lines, missed lines, and missed/extra lines relative to the expert oncologist reference. The * denotes that the research coordinators were compared against the oncologist reference censored at June 1, 2022 (681 of 716 lines); the 35 lines that began after that date were excluded for all comparisons and calculations.

| Line-level error metric | First oncologist | Second oncologist | Gemini 2.5 Pro | GPT-5 | Research coordinator |
| --- | --- | --- | --- | --- | --- |
| Total lines | 716* | 704 | 740 | 714 | 689* |
| Temporally matched lines (start ±90 d) | - | 668 | 662 | 632 | 592 |
| Reference lines without a temporal match, per patient | - | 0.5 | 0.5 | 0.8 | 0.9 |
| Predicted lines without a temporal match, per patient | - | 0.4 | 0.8 | 0.8 | 1.0 |
| Patients with matching line count | - | 68 | 46 | 52 | 44 |

Research coordinators performed worse than all LLMs on systemic therapy abstraction, with a patient-level Jaccard similarity of 0.75 (95% CI, 0.72 to 0.79) for prescribed anti-cancer drugs and 0.49 (95% CI, 0.44 to 0.54) for the lines of therapy. Even after we post hoc collapsed all aromatase inhibitors and taxanes into single entities, research coordinators still had worse performance than the worst LLM for line reconstruction (0.60 vs 0.64). Qualitatively, abstraction performance was stable for the best-performing two LLMs across large and long charts, but performance declined for research coordinators as the chart got bigger (Supplementary Figure 5). A representative treatment timeline for a complex patient is shown in Supplementary Figure 6.

To explore why there were differences between the oncologists and the LLMs, we manually reviewed a random sample of 360 abstraction instances from 20 patients for the two oncologists and the four LLMs. Jaccard similarity and F1 scores were similar in this subset compared with the overall cohort (Supplementary Table 7). For the oncologists, Gemini 2.5 Pro, and GPT-5, most discrepancies reflected judgment calls: how to split or merge therapies into individual lines. Missed events were often short courses of therapy such as a few weeks of endocrine therapy, though adjuvant chemotherapy was missed by one LLM for one patient. The older models (GPT-4o and DeepSeek R1) had qualitatively different failure modes. While differences in line splits and merges were present, the older models also had response truncation, unsupported drug insertion, internally inconsistent dates, and chronology errors. A full qualitative review and comparison is presented in Supplementary Table 3 for non-therapy abstractions and Supplementary Table 8 for therapy lines.

### Comparison of survival and effect estimates

Cohort-level survival estimates were similar when LLM-derived datasets were used in place of expert-derived datasets (Figure 4). Because fewer than half of patients had a documented date of death, overall survival estimates were sensitive to censoring assumptions, with the median overall survival changing by 3 years depending on the censoring rule used. Censoring rules were applied identically to both data sets and did not change the similarity of survival estimates. Median overall survival was 78.2 months for the expert and 78.2 months for the two LLMs (log-rank P = 1.00 and 0.99). Median recurrence-free survival was 34.9 months for the expert and 35.4 and 35.2 months for the two LLMs (P = 0.94 and 0.88) and had much less missingness meaning that survival estimates were similar regardless of censoring choice (Figure 4).

**Figure 4.**
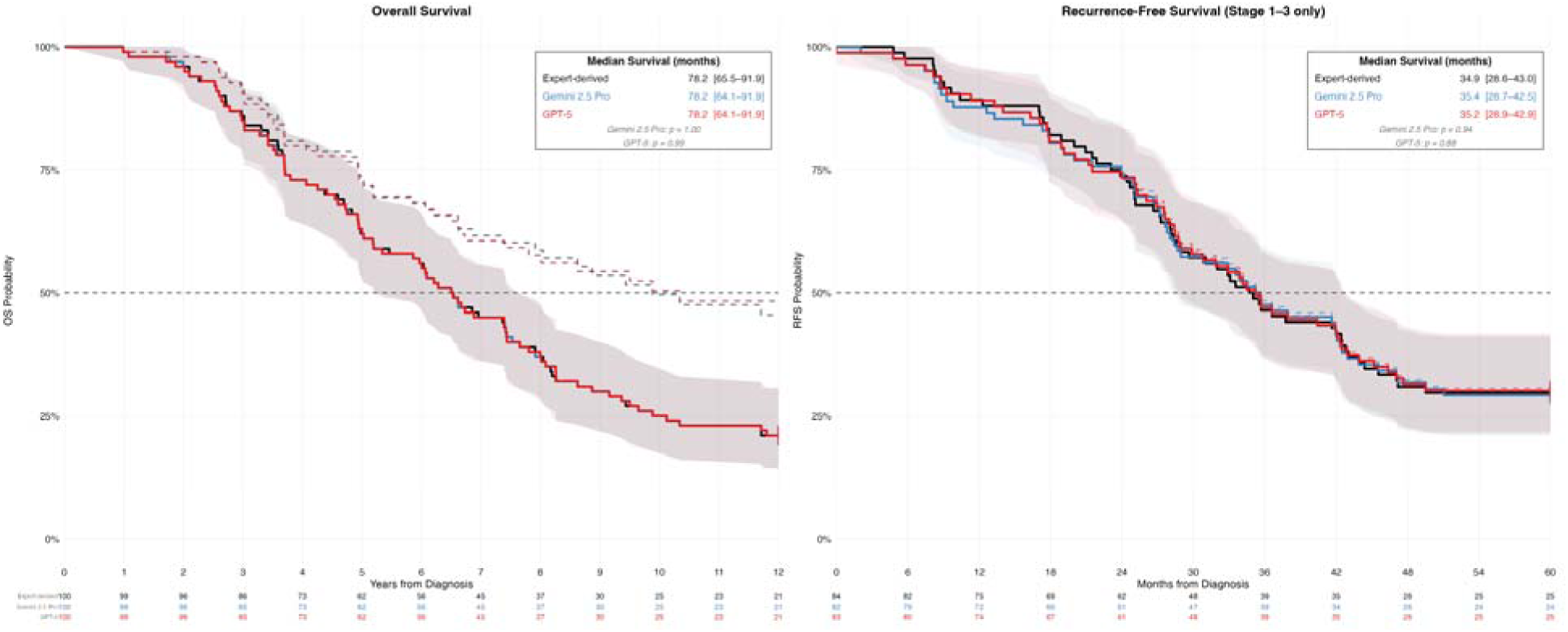
Survival estimates derived from expert vs LLM chart abstraction. Kaplan–Meier curves comparing survival estimates generated from expert-derived data and LLM-derived data for the two best-performing LLMs (Gemini 2.5 Pro and GPT-5). Differences between expert and LLM curves were separately assessed using log-rank tests. The solid and dashed curves are an application of symmetric censoring rules: the solid lines assume death at last follow-up while the dashed lines are censoring of the charts unless there is a documented date of death. This is displayed to show the effect of different analytic assumptions on the end figures regardless of abstractor. (A) Overall survival. (B) Invasive recurrence-free survival among patients with stage 1 – 3 diseases.

When comparing the hazard ratio and confidence intervals for the risk of death for disease stage they were similar for expert-derived and LLM derived datasets, though small numbers led to large confidence intervals: 2.67 (95% CI, 1.26 to 5.63) in the expert-derived dataset, compared with 2.58 (95% CI, 1.25 to 5.32) and 2.85 (95% CI, 1.38 to 5.86) for the best two LLMs. For hormone receptor-negative vs hormone receptor-positive disease, the hazard ratio was 2.06 (95% CI, 1.14 to 3.73) in the expert-derived dataset, compared with lower estimates that did cross 1.00 for the LLMs with ratios of 1.60 (95% CI, 0.87 to 2.91) and 1.66 (95% CI, 0.91 to 3.00) in the corresponding LLM-derived datasets. Cochran Q heterogeneity testing did not identify significant differences in the estimates (Supplementary Table 9).

### Validation in an Independent Clinical Cohort of Young Breast Cancer Patients

In an independent cohort of 97 young patients at Stanford with hormone receptor–positive invasive breast cancer diagnosed before pregnancy, expert manual abstraction identified 11 recurrences, compared with 13 and 13 identified by the two leading LLMs. One discordant case was an expert error; the other was an LLM error. The LLM error occurred in the smallest chart in the cohort—the recurrence event was identified through outside hospital notes and occurred in a 7-year documentation gap in the Stanford medical record. Receipt of adjuvant endocrine therapy had an F1 score of 0.99 and 0.96 for the two LLMs.

## Discussion

In this retrospective study of 100 patients with complex breast cancer histories, a retrieval pipeline using general-purpose LLMs abstracted a broad range of clinical variables from the medical record with high concordance with expert oncologists. Concordance was highest for variables anchored to pathology, such as estrogen receptor status, and lowest for variables requiring clinical reasoning, such as why a treatment was stopped. For anti-cancer drug abstraction, the best-performing model approached inter-oncologist agreement, while exact therapy-line reconstruction for the LLMs remained significantly lower than the two oncologists compared to each other. Despite differences at the individual patient level, cohort level survival and hazard ratio estimates were similar when LLM-derived datasets were compared to expert-derived datasets.

Manual chart abstraction is a major bottleneck in clinical research. Because we write notes in unstructured text, answering many of the clinically meaningful questions is impractical because creating the analytic dataset is too labor-intensive.^5^ We have high uncertainty for many clinical questions when, in principle, the information needed to answer these questions exists in the medical record. This is especially true for rare and complex diseases, settings with heterogeneous practice patterns, areas in which unreplicated retrospective studies dominate clinical decision-making, and domains with less mature disease registries than cancer. One potential implication of this approach, if validated across institutions and diseases, is that sites could use a common abstraction pipeline while keeping patient-level data local.^13^ This could make it easier for multi-institutional consortia to build larger, harmonized cohorts to address a broad range of clinically important questions.

Our results highlight challenges that extend beyond LLMs to any research in which chart review is a central component. Sparse documentation remains a major barrier, particularly in fragmented health systems such as the United States. In health systems with more centralized longitudinal records, LLM-based (and human) abstraction may perform better because more of the relevant clinical history is available and less imputation is needed for missing data. In our study, the lack of high-quality death records illustrates how missing data and analytic assumptions can materially shift results: for overall survival, changing the censor rule changed the survival estimate by 3 years. In contrast, for recurrence we had low missingness and so the estimates, regardless of censoring, had minimal difference (Figure 4). Variables, syndromes, and diseases with ambiguous clinical definitions also remain difficult to abstract and study. Poorly designed abstraction frameworks, whether applied by an LLM or a human abstractor, will generate poor datasets.^16^ The expertise of the abstractor matters as seen in the gap between clinical oncologists and the research coordinators. There was also a gap between the older and newer models used: in the audited line-of-therapies, Gemini 2.5 Pro and GPT-5 disagreements primarily involved how to group drugs into lines, whereas the older models (GPT-4o and DeepSeek-R1) also had chronology errors, truncation, and insertion of an unsupported drug. These more concerning errors were not seen in non-drug tasks (Supplementary Table 1) which were considered simpler tasks than reconstructing the lines of therapy (Supplementary Table 3.)

As LLM-based abstraction becomes more common, whether through abstractors copying and pasting into chatbots or through validated pipelines, it will be critical to establish norms for study design, transparent definitions, statistical rigor, and benchmarking against domain experts.^17^ Most retrospective studies are fundamentally correlative and, on their own, do not support causal inference.^18^ Scaling abstraction will not get us closer to the truth without corresponding scientific rigor.

Over the past two decades, many studies have used natural language processing (NLP) to structure information from clinical notes and reports. Since the LLM revolution, most newer approaches have either focused on narrowly selected inputs, often because of context-window limits, or relied on substantial task-specific systems.^19^ Huang et al., for example, used prompt-engineered general-purpose LLMs to extract structured data from individual pathology reports. This approach requires identifying and providing the relevant document in advance.^20^ Our goal was to remove that manual document-selection step and instead use a single pipeline that could use the whole record for many different questions. Another major branch of the literature uses fine-tuned or foundation models, which can be powerful tools but need specialized infrastructure to develop.^21^ Gupta et al. and Jee et al. relied on fine-tuned or task-specific models and substantial annotation and technical development; Jee et al., for example, combined manually curated clinical labels, multiple NLP models, and institution-specific genomic data across thousands of patients at Memorial Sloan Kettering.^11,13^ Tariq et al. is the closest comparator to our treatment-line analysis, using a hybrid parser and a fine-tuned LLM to reconstruct breast cancer treatment pathways from longitudinal clinical notes.^22,23^ Our study was conducted on a laptop as the goal was to limit the need for specialized on premises compute infrastructure. Direct performance comparisons between these studies are limited because they use different patient populations, diseases, institutions, and retrieval methods. Our study complements this literature by asking how far relatively common infrastructure—access to patient records, a secure computing environment, and API access to a general-purpose LLM—can go.

This study has several limitations. It was conducted in a single healthcare system and focused on a single disease, and external validation is needed to determine whether performance generalizes across institutions, patient populations, EHR environments, and diseases. Technical portability and performance generalizability are distinct: although the pipeline can ingest data exported from different systems, local mapping and validation would still be required to use the pipeline even in identical populations. Effective abstraction was contingent on the relevant data being present in the record. Errors could result from missing data, imperfect retrieval, poorly designed prompts or abstraction definitions, or LLM reasoning errors. Older LLMs were more prone to concerning errors such as insertion of erroneous therapies compared to the newest LLMs. For most non-systemic variables, the primary comparator was a single oncologist abstraction with targeted rather than random dual review, so there could be errors by the oncologists. Because the research-coordinator abstraction was performed in 2022, the comparison covers most but not all oncologist-abstracted lines: 35 oncologist abstracted lines (5%) occurred after June 1, 2022, and therefore is a limitation in direct research coordinator to LLM comparisons. Simplifying variables for analysis can itself lose clinically meaningful information, as we collapsed the reasons for treatment discontinuation into progression vs non-progression.

Within these limitations, these findings support the feasibility of using LLM-based abstraction to build retrospective datasets that would otherwise be difficult to assemble manually. Realizing this potential will require carefully designed abstraction tasks, validated outputs, and external validation across institutions and clinical settings.

## Methods

### Study design and objectives

We compared a fully automated retrieval-based pipeline using commercially available general-purpose LLMs vs expert oncologist chart review for a range of variables (Table 4). The primary endpoints were agreement between LLM-derived and expert-derived data on cancer recurrence, including whether recurrence occurred and its timing, and on the number and composition of systemic treatment lines. Secondary endpoints were agreement on other clinical variables, comparison with research coordinators for selected variables, inter-oncologist variability in systemic therapy abstraction, and preservation of downstream inference when LLM-derived datasets were substituted for expert-derived datasets. This study was approved by the Stanford University Institutional Review Board under protocol 19482. The requirement for informed consent was waived by the Institutional Review Board because the study involved retrospective analysis of existing clinical data. All research was conducted in accordance with the Declaration of Helsinki and Stanford’s institutional guidelines and regulations. The reporting framework outlined in TRIPOD-LLM was used to guide this manuscript.^24^

**Table 4.** Clinical variables extracted from the electronic medical record and evaluation metrics. Date-based variables were evaluated using concordance within ± 90 days. Categorical variables were evaluated using exact concordance. Systemic therapy abstraction included both exact lines of therapy and component anti-cancer drugs, as well as matched-line start and stop dates, treatment intent, and reason for discontinuation. For genetic testing we had access to the files from Foundation Medicine and from the companies doing the germline genetic testing and so this was used as the reference rather than oncologist abstraction. The ^ denotes that patient-level mean Jaccard similarity was used as the benchmark for inter-expert agreement in systemic therapy abstraction. The # denotes that for analysis, discontinuation reasons were collapsed to disease progression vs other. Progression included documented progressive disease, hospice enrollment related to advancing cancer, death, or decline in performance status in the setting of progressing disease. Other included planned completion, toxicity or adverse event, patient preference, clinician choice, and transfer of care.

|  | Comparison | Evaluation metric |
| --- | --- | --- |
| **Timing of key events** |  |  |
| **Date of birth** | Structured record vs LLM | Concordance within $\pm 90$ days |
| Date of diagnosis | Oncologist vs LLM | Concordance within $\pm 90$ days |
| Date of first locoregional recurrence | Oncologist vs LLM | Concordance within $\pm 90$ days |
| Date of first metastatic diagnosis | Oncologist vs LLM | Concordance within $\pm 90$ days |
| Documented death date identified | Oncologist vs LLM | Agreement on presence of a documented death date |
| Recurrence occurred (yes/no) | Oncologist vs LLM | Exact concordance |
| **Clinical information** |  |  |
| Hormone receptor status | Oncologist vs LLM | Exact concordance |
| HER2 status | Oncologist vs LLM | Exact concordance |
| Original stage | Oncologist vs LLM | Exact concordance |
| Primary treating oncologist | Oncologist vs LLM | Exact concordance |
| **Genetic testing** |  |  |
| Germline BRCA1 or BRCA2 pathogenic variant identified? | Genetics clinic database vs LLM | Mutated vs not mutated / not found |
| Somatic PIK3CA mutation present? | Genetics clinic database vs LLM | Mutated vs not mutated / not found |
| Somatic ESR1 mutation present? | Genetics clinic database vs LLM | Mutated vs not mutated / not found |
| **Systemic therapy** |  |  |
| All prescribed anti-cancer drugs | Second oncologist<sup>^</sup> vs LLM vs research coordinator | Precision, recall, F1 score, fraction of drugs identified |
| Exact lines of therapy | Second oncologist<sup>^</sup> vs LLM vs research coordinator | Patient-level Jaccard similarity; line-level precision, recall, and F1 score |
| Start and stop date of each matched treatment line | Second oncologist vs LLM vs research coordinator | Concordance within $\pm 30$ days for matched lines |
| Reason for discontinuation (progression vs other)<sup>#</sup> | Second oncologist vs LLM vs research coordinator | Exact concordance for matched lines |
| Treatment intent (curative vs palliative) | Second oncologist vs LLM vs research coordinator | Exact concordance for matched lines |

### Study cohort and data extraction

This was a descriptive proof-of-concept study, and no formal sample-size, power, or precision calculation was performed. A target sample of 100 patients was selected based on the feasibility of conducting detailed manual review of the records. The sampling frame was selected to enrich for complex disease trajectories while retaining variation in documentation patterns and treating clinicians across the academic flagship campus and a community-affiliate practice. Therefore, we used an institutional breast cancer cohort enriched for complex clinical trajectories and treatment histories who had undergone either tissue or blood-based FoundationOne testing (n=206) (Foundation Medicine, Cambridge, MA, USA).^25,26^ From this cohort, 100 patients were randomly selected using Python’s random.sample() function.

### Reference standard generation and human comparators

Four medical oncologists specializing in breast cancer care (JCD, NHD, MBM, and JLC) were each assigned a random subset of patients and asked to abstract the variables listed in Table 4. Experts were blinded to LLM results. They were instructed to use all available data in the medical record, excluding external notes accessible only through Epic’s external note viewer Care Everywhere (Epic Systems, Verona, WI, USA).^27^ Experts were provided the clinical definitions used in the study (Supplementary Table 10), but were not told how to abstract the variables.

For most clinical variables, a single expert abstraction served as the reference standard. Because systemic therapy abstraction was expected to be the most difficult task, and the hardest to directly measure, a second oncologist independently abstracted the same systemic therapy variables for each patient. This allowed inter-expert variability, rather than a single abstraction, to serve as the benchmark for performance. Experts were asked to reconstruct lines of therapy, list their component drugs, assign start and stop dates, classify treatment intent as curative or palliative, and determine whether discontinuation was due to disease progression or another reason such as toxicity or patient preference.

As an additional point of comparison, research coordinators had previously abstracted the original date of diagnosis, the date of distant metastatic disease, and the same systemic therapy variables. Because the research coordinator abstraction was performed in 2022, all comparisons involving research coordinators were restricted to clinical information available before June 1, 2022.

### LLM abstraction pipeline overview

On January 10, 2026, we downloaded all available text-based electronic medical record documents for each patient from Stanford’s research warehouse. Captured documents spanned from Stanford’s adoption of Epic in 2008 to January 9, 2026. At the time of the study, data from Epic were exported to Stanford’s research data warehouse which is hosted on Google Cloud.^28^ Pathology reports, demographics, clinical notes, and medication administration records were exported as separate CSV files, with one row per document and one folder per patient. Original metadata, including document type, author, and entry date, were preserved. The warehouse does not preserve images, scanned documents, or screenshots in the notes. No manual annotation, normalization, cleaning, or editing of the source documents was performed before ingestion into the pipeline.

The pipeline takes each narrative document and segments it into roughly paragraph length ‘chunks’ of text. These chunks then can be retrieved and passed to the LLM with a task-specific prompt. Each patient chart was embedded into its own vector space to avoid cross-patient contamination. We used a locally hosted biomedical embedding model, NeuML_bioclinical-modernbert-base-embeddings with embedding dimensionality of 768.^29^ All text-containing fields were segmented into chunks of 1,024 tokens with 128-token overlap. We chose this chunk size arbitrarily with the idea that this would provide enough context for the ID statements and treatment histories for the patients.

For each variable in Table 4, retrieval was restricted to the document types most relevant to manual abstraction of that variable, and custom prompts were used for each variable. For example, for identifying the original date of cancer diagnosis the pipeline first sends pathology report chunks to the LLM and then a separate prompt sends clinical notes. Retrieval used a combination of search strategies to find candidate text chunks including exact word search, BM25, and semantic embedding queries over an hnswlib index, depending on the variable (Supplementary Table 11).^30,31^ Retrieval parameters were iteratively set during pilot development and then fixed before final evaluation.

Retrieved text chunks were then provided to commercially available LLMs through Stanford’s HIPAA-compliant SecureGPT environment. The pipeline stamped the chunks cited or used during generation, permitting post hoc analysis of the retrieval rank of evidence used for each task. Model versions were fixed at the time of extraction and included GPT-5 (gpt-5-2025-08-07), GPT-4o (gpt-4o-2024-11-20), DeepSeek-R1 (DeepSeek-R1-0528), and Gemini 2.5 Pro (gemini-2.5-pro), which were available through the Stanford SecureGPT interface.^32^ We chose the newest models available at the time, along with a pair of older models to examine performance changes as frontier models improve with time. All models were run with deterministic settings (temperature = 0), except for gpt-5 where this cannot be selected, and all data stayed within Stanford’s secure institutional computing environment. We used the max tokens for the model for all tasks; no seed was set and top p was at its default. Reported performance is for the integrated system and not the LLMs in isolation.

Tasks were stateless (i.e. no chat history), but for multi-pass tasks, intermediate outputs were carried forward. After pilot development using charts from the cohort, prompts, retrieval parameters, and task-specific workflows were fixed before final model evaluation. For a given task, all models received identical text chunks and prompts. The LLMs had no access to expert annotations nor structured reference data. The embedding and all experiments were run on a 2024 MacBook with an M4 Pro processor and 48 GB of RAM. LLM inference was done through the approved SecureGPT API endpoint.

### Outcome Measures

The primary endpoints of this descriptive proof-of-concept study were agreement between LLM-derived and expert-derived abstraction for recurrence dates and lines of systemic therapy. We also evaluated the concordance for the additional variables listed in Table 4. Supplementary Table 10 shows the definitions used for abstraction. For all non-drug variables, we prespecified greater than 90% agreement with expert-derived data for at least two LLMs as an arbitrary threshold for encouraging performance. For date-based variables, agreement was defined as concordance within ± 90 days, which we prespecified as a clinically meaningful threshold. Categorical variables required exact concordance. For mutation testing, the primary analysis focused on mutation presence, with all other outcomes treated as negative or not found.

For prescribed systemic therapy, we evaluated both exact lines of therapy and the anti-cancer drugs contained within those lines. We prespecified inter-oncologist variability as the benchmark for systemic therapy performance. We considered overlap of the confidence intervals for patient-level mean Jaccard similarity between at least two LLMs and the second oncologist as encouraging performance.^33^ After therapy-name normalization, predicted and reference lines were matched using an optimal, order-preserving one-to-one assignment based on temporal proximity. If a line had no start date, then it was matched on identical regimen composition alone. We also calculated precision, recall, and F1 score for both line-level and drug-level extraction. Additional analyses included agreement on the number of therapy lines and, among matched lines, agreement on start and stop dates within ± 30 days, treatment intent, and reason for discontinuation.

### Statistical Analyses

After abstraction, we created two separate analytic datasets: one LLM-derived and one expert-derived. We then examined overall survival from initial breast cancer diagnosis and invasive recurrence-free survival.

These survival estimates were measured using Kaplan-Meier methods applied symmetrically to the LLM and human data. In Figure 4, we present two censoring methods: if there was no date of death recorded, the primary analysis uses date of last follow-up as death; the dashed lines use the conventional censoring for these patients.^34^ The cox proportional-hazards models use the conventional censoring at the last documented follow-up date. To compare effect estimates in human-derived and LLM-derived datasets, we evaluated univariate hazard ratio estimates for stage (4 vs 1 – 3) and hormone receptor status (negative vs positive) on overall survival. We separately looked at stage (3 vs 1 – 2) and hormone receptor status on recurrence-free survival. We compared hazard ratio estimates and 95% confidence intervals from expert-derived and LLM-derived datasets and used Cochran’s Q test as a test for heterogeneity between effect estimates.

Confidence intervals for all agreement and performance metrics were estimated using bias-corrected and accelerated bootstrap resampling at the patient level with 1,000 iterations; proportions at 0% or 100% used Wilson score intervals.^35^ All analyses were conducted in R version 4.5.2 (R Foundation for Statistical Computing, Vienna, Austria) using base functions and the *boot* and *survival* packages. The analytic code is available on <u>GitHub</u>.

### Validation in an Independent Clinical Cohort of Young Breast Cancer Patients

To examine performance in a clinically distinct population, we evaluated a previously manually abstracted cohort of young patients diagnosed with breast cancer before becoming pregnant.^36^ We compared recurrence detection and receipt of adjuvant endocrine therapy for patients who had an initial invasive diagnosis and hormone receptor–positive disease. We compared single abstraction by breast oncologists to the two best-performing LLMs. The pipeline, retrieval strategy, and prompts were applied to this cohort without modification.

### Audits and Error Analysis

To characterize line-of-therapy errors, we randomly selected 20 patients using Python’s random.sample() function and manually reviewed the first three lines of therapy for the two clinicians and four LLMs. We qualitatively evaluated differences in the quality of the abstraction and the error patterns. We compared F1 and Jaccard performance in this subset with the overall cohort to assess whether model performance was similar in the reviewed sample.

For non-systemic clinical variables, we performed an additional discordance audit when at least two LLMs agreed with one another and disagreed with the primary expert abstraction. These cases were reviewed by at least two clinicians and adjudicated by consensus to establish the final reference label. For the qualitative error analysis, we reviewed all instances for the best two models where they were discordant with the reference label.

## Declaration Statements

### Data Availability

The patient-level clinical records and derived patient-level annotations generated and analyzed during this study are not publicly available because they contain protected health information and are subject to institutional privacy and regulatory restrictions. Access may be considered for qualified researchers upon reasonable request to the corresponding author, subject to institutional review and approval, applicable institutional review board authorization, and execution of an appropriate data use agreement.

### Code Availability

The study-specific analytic code used to generate the results reported in this manuscript is publicly available at https://github.com/MrJimb0/junior_first_paper_analytic_code. The source code for the automated chart-review pipeline is publicly available at https://github.com/MrJimb0/junior.

## Acknowledgements

We would like to thank Jessica Orford, Sonia Rios-Ventura, and Rozelle Laquindanum for their contributions as research coordinators. This work was supported by the Stanford Center for Digital Health. The funder had no role in the study design; data collection, analysis, or interpretation; preparation of the manuscript; or the decision to submit the manuscript for publication.

## Author Contributions

J.C.D. conceived the study, developed the methodology, curated and analyzed the data, created the visualizations, administered the project, and drafted the manuscript. M.B.M., M.S., and N.H.D. curated the data and reviewed and edited the manuscript. M.B.R. contributed to the methodology and formal analysis and reviewed and edited the manuscript. C.C. curated the data and reviewed and edited the manuscript. A.W.K. and J.L.C.-J. contributed to the study conception, methodology, formal analysis, and data curation and reviewed and edited the manuscript. All authors reviewed and approved the final manuscript.

## Competing Interests

James C. Dickerson reports stock ownership in Johnson & Johnson and Merck. Jennifer L. Caswell-Jin reports research funding paid to her institution from eFFECTOR Therapeutics and Novartis. Christina Curtis reports advisory or consulting relationships with AstraZeneca, Bristol Myers Squibb, Pfizer, Genentech, DeepCell, and 3T Biosciences, as well as stock ownership in DeepCell and 3T Biosciences. The other authors do not have a competing interest.

## Prior Presentations

This work was presented as a poster at American Society of Clinical Oncology Annual Meeting 2025 and the San Antonio Breast Cancer Symposium 2025

## Notes

### Competing Interest Statement

JCD: Stock Ownership: Johnson & Johnson, Merck. JLC: research funding to her institution from Effector Therapeutics and Novartis.

### Author Declarations

Our study was approved by the Stanford IRB under protocol 19482

### Summary of Updates

Only changes were to update the funder to the Center for Digital Health and to add Dr. Curtis who provided the linkage to the genetic information and provided assistance with further revisions of the manuscript

