## Appendix for "Abstraction of Longitudinal Breast Oncology Records Using a Retrieval Pipeline with General-Purpose Large Language Models"

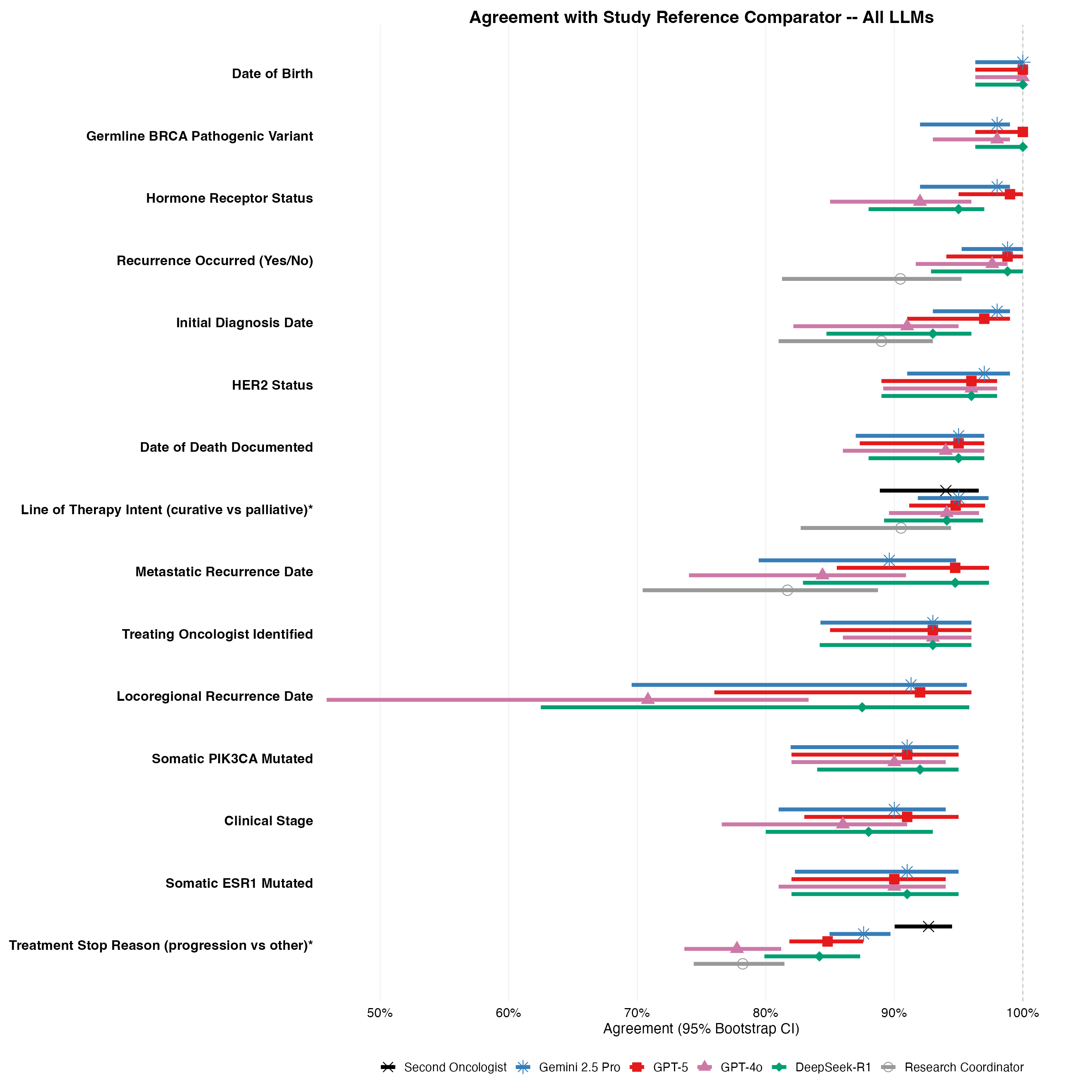

**Supplementary Figure 1. Concordance with oncologist abstraction across all evaluated LLMs.**
Expanded Forest plot showing agreement with expert oncologist abstraction and 95% bootstrap confidence intervals for all evaluated LLMs, the second oncologist, and research coordinators across the clinical variables shown in Figure 2. For locoregional recurrence date, agreement was evaluated only among patients in whom both the expert and the comparator identified a locoregional recurrence date; missed and false-positive events were assessed separately. For metastatic diagnosis date, patients with *de novo* metastatic disease were excluded. Germline BRCA1/2, somatic PIK3CA, and somatic ESR1 were analyzed as binary mutation present or not. Reference comparators were linked genetic testing records for germline and PIK3CA mutations and the oncologist for all others variables. The * denotes that the treatment intent and reason for discontinuation were evaluated only among matched therapy lines

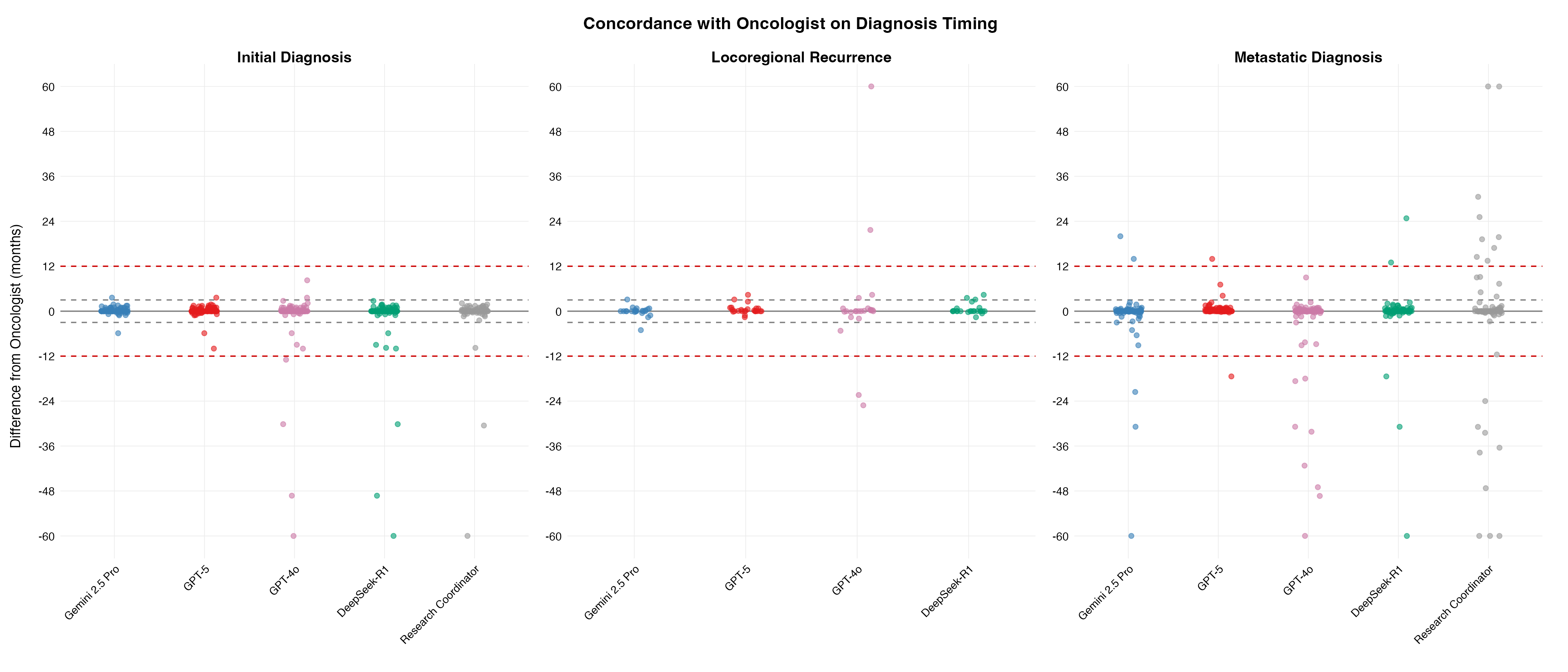

**Supplementary Figure 2. Dispersion of event dates relative to expert oncologist review.**
Scatterplots showing the difference in months between LLM-derived and oncologist-derived dates for initial diagnosis, locoregional recurrence, and metastatic diagnosis. Values are calculated as comparator minus expert. Positive values are when the LLM assigned a later date than the expert and the reverse for negative dates.

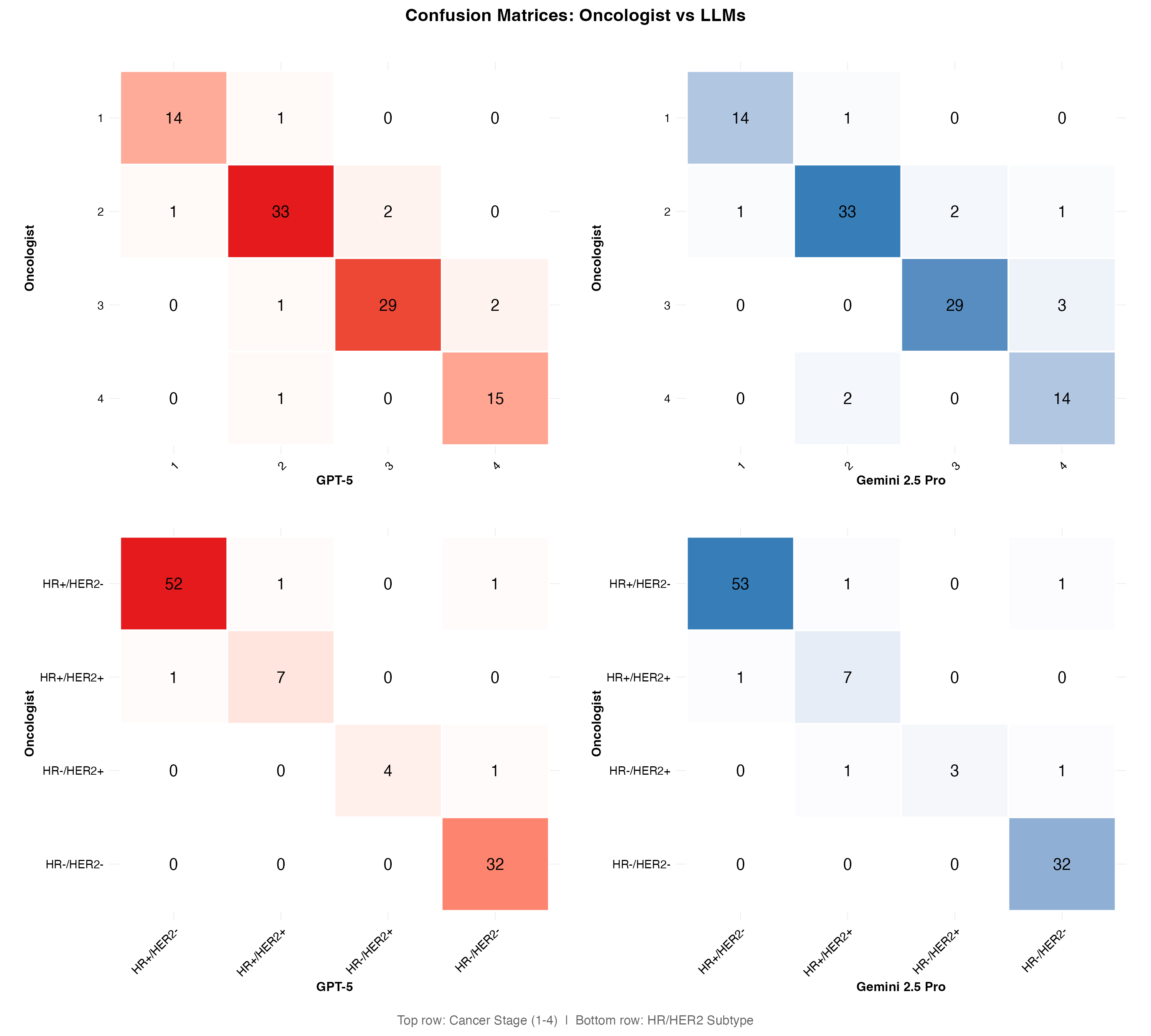
**Supplementary Figure 3. Confusion matrices for stage and HR/HER2 subtype.**
Confusion matrices comparing expert oncologist abstraction with the two best-performing LLMs for clinical stage and HR/HER2 subtype.

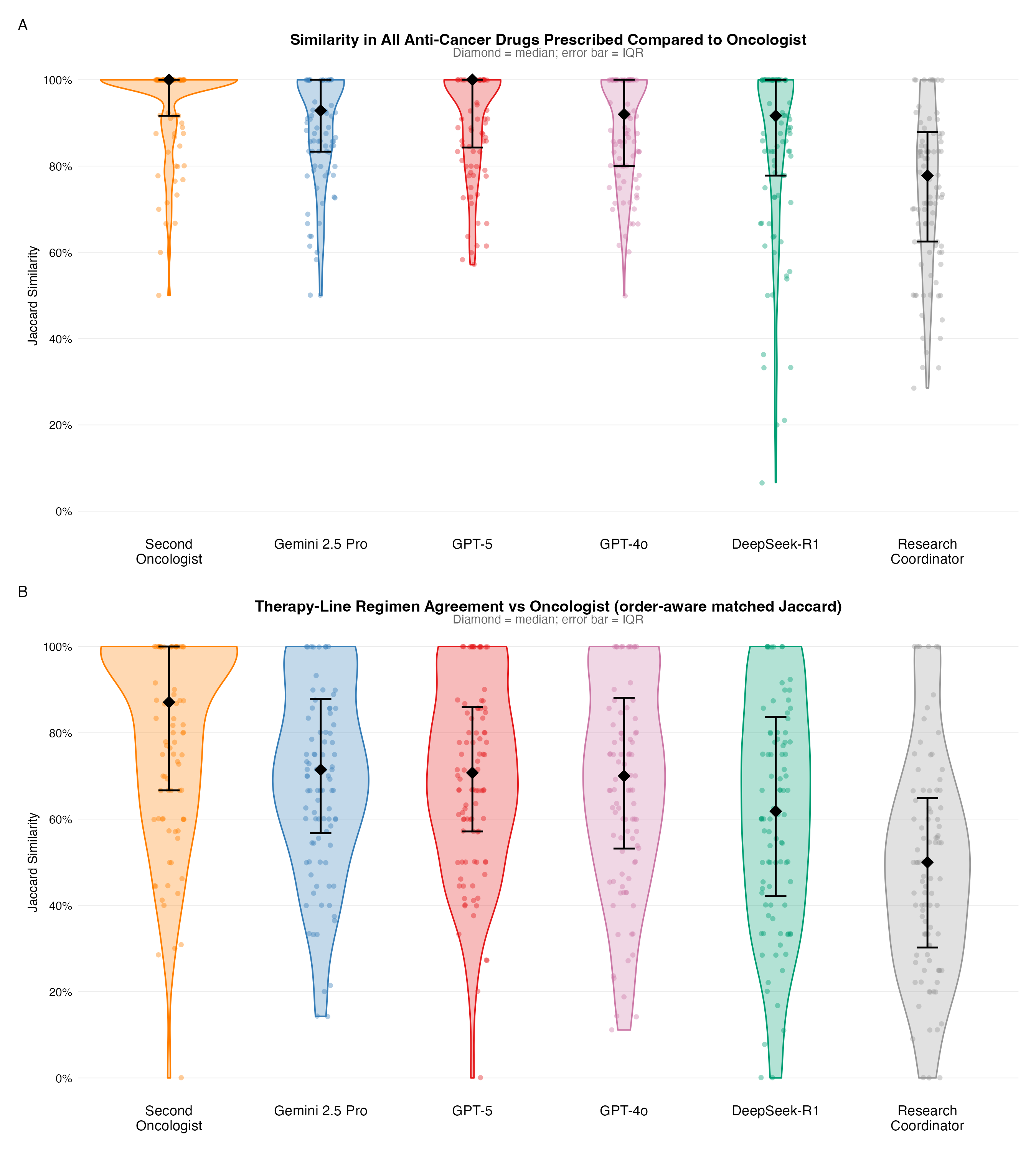

**Supplementary Figure 4. All LLM results for the drug and line Jaccard similarity**

Violin plots showing patient-level Jaccard similarity for abstraction of all prescribed anti-cancer drugs (Panel A) and therapy lines (Panel B). Diamonds are at the medians and bars are the interquartile ranges.

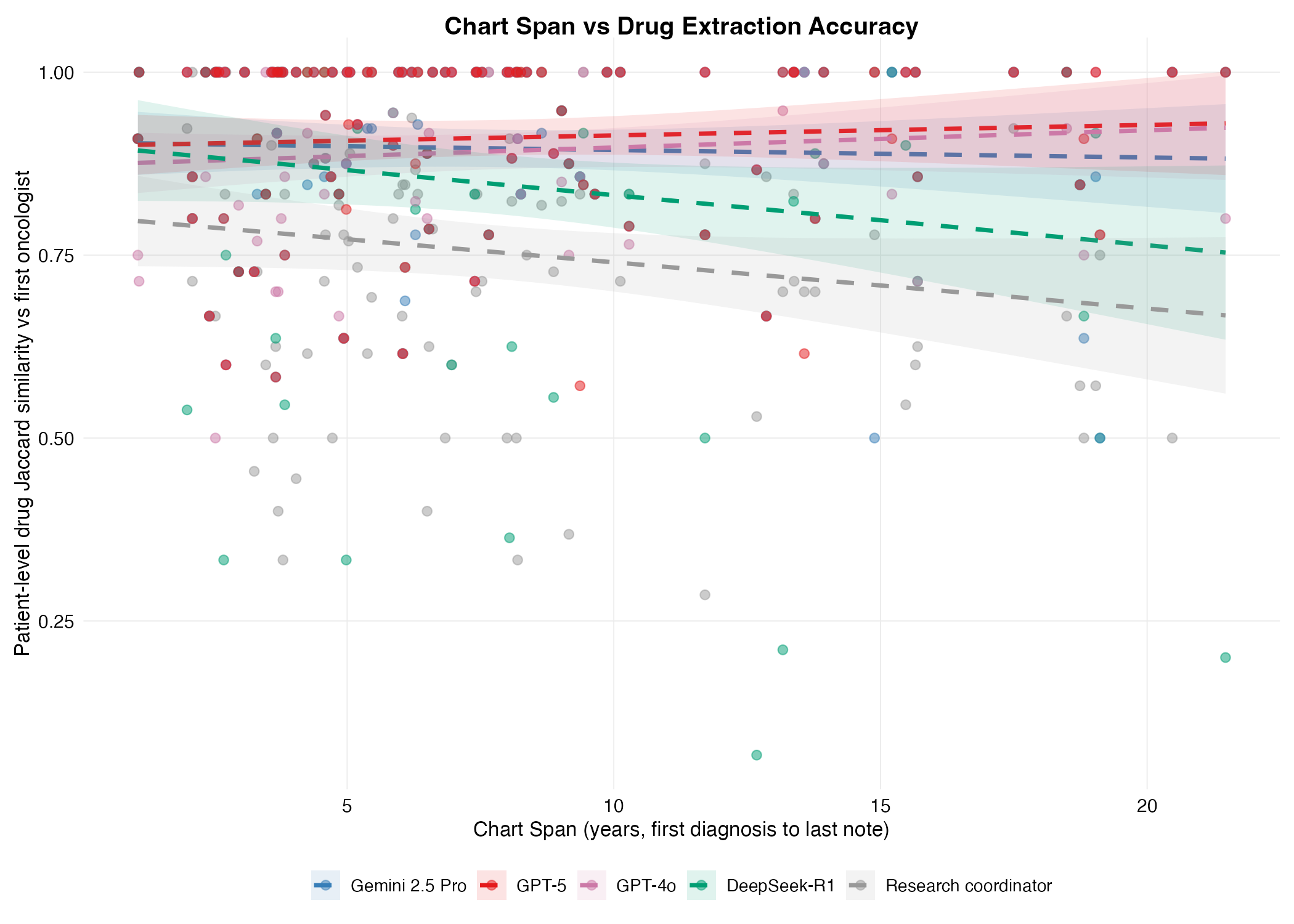

**Supplementary Figure 5. Chart span and drug extraction accuracy.**
Association between duration of documented clinical care and patient-level Jaccard similarity for anti-cancer drug extraction. Qualitatively, scores dropped for the longest charts for the research coordinators and DeepSeek-R1 but drop off for large charts was less noticeable for the other models.

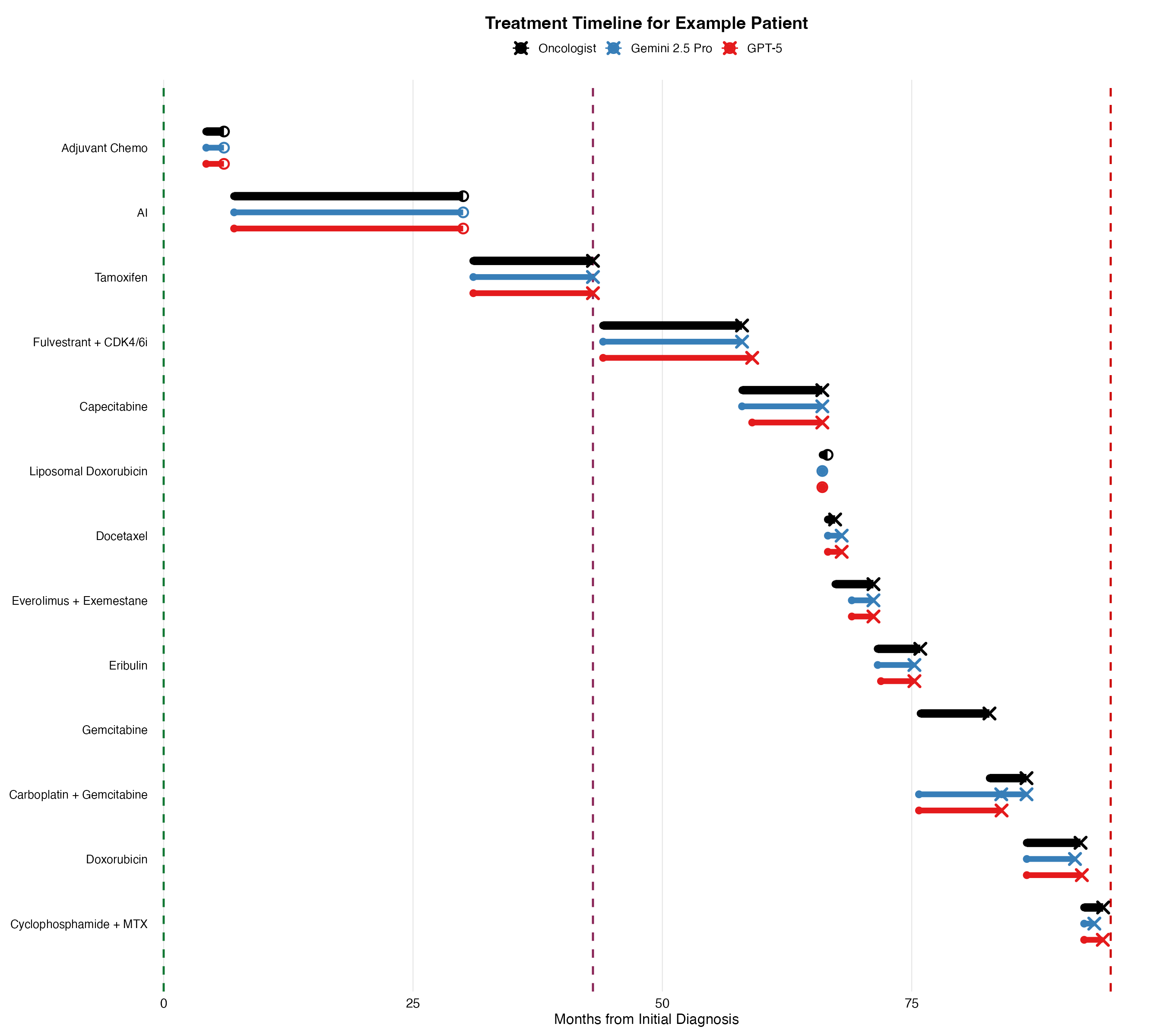

**Supplementary Figure 6. Representative reconstructed patient timeline.**
Example longitudinal timeline reconstructed from the medical record. The first vertical line is the original diagnosis, the second the date of metastatic recurrence, and the final line is the date of death (oncologist and LLM dates overlap closely). An X represents stoppage due to progression; an O represents stoppage due to an adverse event.

| **Clinical domain / variable** | **Second oncologist** | **Gemini 2.5 Pro** | **GPT-5** | **GPT-4o** | **DeepSeek-R1** | **Research coordinator** |
| --- | --- | --- | --- | --- | --- | --- |
| **Timing of key events** |  |  |  |  |  |  |
| Documented death date identified | — | 95%  [88 - 97] | 95%  [88 - 98] | 94%  [86 - 97] | 95%  [87 - 98] | — |
| Date of birth (±90 d) | — | 100%  [96 - 100] | 100%  [96 - 100] | 100%  [96 - 100] | 100%  [96 - 100] | — |
| Date of diagnosis (±90 d) | — | 98%  [92 - 99] | 97%  [91 - 99] | 91%  [81 - 95] | 93%  [84 - 96] | 97%  [90 - 99] |
| Recurrence occurred (yes/no) | — | 99%  [93 - 100] | 99%  [95 - 100] | 98%  [89 - 99] | 99%  [93 - 100] | — |
| Date of first locoregional recurrence (±90 d) | — | 91%  [70 - 96] | 92%  [68 - 96] | 71%  [46 - 83] | 88%  [62 - 92] | — |
| Date of first metastatic diagnosis (±90 d) | — | 90%  [81 - 95] | 96%  [89 - 98] | 86%  [76 - 91] | 95%  [86 - 98] | 73%  [63 - 80] |
| **Clinical information** |  |  |  |  |  |  |
| Original HR/HER2 subtype | — | 95%  [88 - 98] | 95%  [87 - 98] | 88%  [79 - 93] | 91%  [82 - 95] | — |
| Hormone receptor status | — | 98%  [91 - 99] | 99%  [95 - 100] | 92%  [85 - 96] | 95%  [85 - 97] | — |
| HER2 status | — | 97%  [89 - 99] | 96%  [88 - 98] | 96%  [89 - 98] | 96%  [88 - 98] | — |
| Original stage | — | 90%  [81 - 94] | 91%  [83 - 95] | 86%  [77 - 91] | 88%  [79 - 93] | — |
| Primary treating oncologist | — | 93%  [86 - 96] | 93%  [86 - 96] | 93%  [86 - 96] | 93%  [85 - 97] | — |
| **Genetic testing** |  |  |  |  |  |  |
| Germline BRCA1/2 mutation identified | — | 98%  [93 - 99] | 100% [96 - 100] | 98%  [93 - 99] | 100% [96 - 100] | — |
| Somatic PIK3CA mutation present | — | 91%  [83 - 95] | 91%  [83 - 95] | 90%  [82 - 94] | 92%  [83 - 96] | — |
| Somatic ESR1 mutation present | — | 91%  [82 - 95] | 90%  [82 - 94] | 90%  [81 - 94] | 91%  [82 - 95] | — |
| **Systemic therapy** |  |  |  |  |  |  |
| All prescribed anti-cancer drugs: Jaccard similarity (mean) | 0.95  [0.92 - 0.96] | 0.90  [0.87 - 0.92] | 0.91  [0.89 - 0.93] | 0.89  [0.86 - 0.92] | 0.85  [0.80 - 0.89] | 0.75  [0.72 - 0.79] |
| All prescribed anti-cancer drugs: precision | 97%  [95 - 98] | 92%  [90 - 94] | 93%  [91 - 95] | 93%  [90 - 95] | 93%  [91 - 95] | 88%  [85 - 90] |
| All prescribed anti-cancer drugs: recall | 98%  [96 - 99] | 97%  [95 - 98] | 97%  [95 - 98] | 96%  [94 - 97] | 90%  [86 - 94] | 83%  [79 - 86] |
| All prescribed anti-cancer drugs: F1 score | 97%  [96 - 98] | 94%  [92 - 95] | 95%  [93 - 96] | 94%  [92 - 95] | 90%  [86 - 93] | 85%  [82 - 87] |
| Systemic therapy lines: regimen Jaccard (mean) | 0.80  [0.76 - 0.84] | 0.70  [0.66 - 0.75] | 0.70  [0.66 - 0.75] | 0.69  [0.64 - 0.73] | 0.62  [0.57 - 0.68] | 0.49  [0.44 - 0.54] |
| Systemic therapy lines: exact-content precision (mean) | 88%  [84 - 90] | 79%  [76 - 83] | 81%  [77 - 84] | 79%  [75 - 82] | 76%  [70 - 79] | 62%  [57 - 67] |
| Systemic therapy lines: exact-content recall (mean) | 87%  [82 - 90] | 82%  [78 - 86] | 81%  [77 - 84] | 80%  [75 - 84] | 73%  [68 - 78] | 63%  [58 - 67] |
| Systemic therapy lines: exact-content F1 (mean) | 87%  [84 - 90] | 80%  [76 - 83] | 80%  [76 - 83] | 79%  [75 - 82] | 73%  [68 - 77] | 62%  [57 - 66] |
| Total predicted therapy lines, No. | 704 | 740 | 714 | 725 | 668 | 689* |
| Number of matched lines | 668 | 662 | 632 | 625 | 575 | 592 |
| Start date of matched treatment line (±30 d) | 98%  [96 - 99] | 94%  [92 - 96] | 96%  [93 - 97] | 91%  [88 - 94] | 90%  [86 - 93] | 95%  [91 - 97] |
| Stop date of matched treatment line (±30 d) | 90%  [86 - 92] | 81%  [76 - 85] | 79%  [73 - 83] | 77%  [72 - 82] | 75%  [70 - 80] | 74%  [69 - 78] |
| Treatment intent for matched lines | 93%  [89 - 96] | 95%  [90 - 97] | 95%  [91 - 97] | 93%  [88 - 96] | 94%  [90 - 97] | 91%  [86 - 94] |
| Reason for discontinuation for matched lines | 92%  [90 - 95] | 87%  [83 - 89] | 83%  [80 - 87] | 77%  [73 - 80] | 85%  [81 - 88] | 78%  [74 - 82] |

**Supplementary Table 1. Full model-by-model abstraction performance across evaluated clinical variables.**
Agreement metrics for all evaluated LLMs across all variables. CIs were estimated using bias-corrected and accelerated bootstrap resampling at the patient level with 1,000 iterations.

The * denotes that the research coordinators were compared against the oncologist reference censored at June 1, 2022 (681 of 716 lines); the 35 lines that began after that date were excluded for all comparisons and calculations.

| **LLM** | **Mean minutes per patient** | **Mean Tokens/Patient** | **Average API Calls Per Patient (Including Repairs)** |
| --- | --- | --- | --- |
| DeepSeek-R1 | 10.9 | 395,923 | 23 |
| Gemini 2.5 Pro | 9.0 | 354,216 | 19 |
| GPT-4o | 1.4 | 280,534 | 18 |
| GPT-5 | 11.3 | 337,304 | 19 |

**Supplementary Table 2. Computational use and runtime of LLM chart abstraction.**

| **Clinical variable** | **Qualitative analysis of errors** |
| --- | --- |
| Date of initial breast cancer diagnosis | The largest outliers were remote diagnoses that required both the expert and the LLM to infer month and/or day from sparse chart context. An example would be similar to a chart stating, “originally diagnosed in 2003,” with no further information. |
| Date of death | In the missed cases, the expert inferred death from context. Instructions for the LLM asked for explicit documentation of death and so it nulled these events. An example would be similar to a nursing note that stated, “spoke with spouse about the patient’s passing last night.” that the expert used as evidence of a death date but the LLMs did not take as an explicit date of death. |
| Date of first locoregional recurrence and metastatic diagnosis | Locoregional dates were often nulled when proceeding metastatic dates. Experts were instructed to document locoregional recurrence preceding metastatic diagnosis, whereas the LLMs often nulled the locoregional recurrence when there was equivocal imaging at that time and then later metastatic confirmation.  Errors that were large (> 1 year difference in expert and LLM) were often from differing weighting of biopsy evidence and clinician documentation. For example, an indeterminate lesion with a negative biopsy. However, the LLM reasoning cited that the clinician, “referred to the patient as metastatic” as the reason for using this as the metastatic date even though the pathology data suggested otherwise.  In another example where the LLMs were > 1 year later than the expert, there was a paragraph in a single non-oncology note stating a recurrence date one year before the first pathologic proof which was the metastatic date the LLMs anchored on. |
| Clinical stage at diagnosis | Most discrepancies were about provenance, and one was due to expert error.  In one case the expert classified as stage 4 because early imaging showed bone disease, but this was ignored briefly clinically and the chart documented “Stage IIA, pT2, pN1mi” disease. These same bone lesions were biopsied shortly thereafter confirming metastatic disease.  In the expert mistake case, the LLMs correctly retained the original pre-neoadjuvant clinical stage whereas the expert used a later restaging note where the clinician had documented the ypTNM stage. This was identified during the audit. |
| Hormone receptor and HER2 status | Errors generally occurred when the original cancer was poorly documented and later pathology reflected a receptor change. For example, no original pathology available for a diagnosis in the early 2000s but notes stating ER-positive where the metastatic recurrent biopsy showed triple-negative disease.  One example where the prompts led to a forced decision was an example of synchronous bilateral primaries with discordant HER2 status forcing the LLMs to pick the left or the right.  In one case, the outside pathology had initially been read as HER2-positive, whereas later biopsies showed HER2-negative. The clinician used the original outside hospital report as evidence of HER2-positive disease for the original diagnosis. |
| Primary treating oncologist | There was one fellow who became faculty and notes written as a fellow were documented as her being the primary author. This meant that when she was a trainee, she was still counted as the treating oncologist despite being supervised by another oncologist that the expert identified as the treating oncologist. The other errors were from patients changing oncologists over time and from sparse outpatient encounters (i.e. second opinion cases treated at other hospital systems). |
| Somatic PIK3CA and ESR1 mutation status | Most false negatives were because of screenshots and pdfs in the media tab being the only places of documentation. However, on audit review, there were cases where the mutation testing was nowhere in the electronic medical record so even a clinician would have reached the same conclusion on chart review. |

**Supplementary Table 3. Qualitative error analysis for non-systemic therapy extraction tasks.**
For each variable we examined the largest outliers for the best two LLMs and then looked for themes or unique cases that could explain the discordant results. Most results for these variables were traceable to differences in interpretation, context, and anchors.

| **Variable** | **LLM** | **Median Chunk Rank** | **% Rank 1** | **% Top 5** | **% > 10** |
| --- | --- | --- | --- | --- | --- |
| Clinical stage | DeepSeek-R1 | 9 | 12% | 33% | 43% |
| Clinical stage | Gemini 2.5 Pro | 9 | 15% | 34% | 44% |
| Clinical stage | GPT-4o | 9 | 14% | 35% | 42% |
| Clinical stage | GPT-5 | 8 | 14% | 36% | 39% |
| Orig diagnosis date | DeepSeek-R1 | 3 | 28% | 63% | 17% |
| Orig diagnosis date | Gemini 2.5 Pro | 4 | 25% | 58% | 21% |
| Orig diagnosis date | GPT-4o | 3 | 31% | 62% | 19% |
| Orig diagnosis date | GPT-5 | 6 | 14% | 49% | 29% |
| Locoreg Rec | DeepSeek-R1 | 5 | 27% | 54% | 19% |
| Locoreg Rec | Gemini 2.5 Pro | 10 | 13% | 35% | 44% |
| Locoreg Rec | GPT-4o | 6 | 28% | 45% | 24% |
| Locoreg Rec | GPT-5 | 10 | 8% | 27% | 50% |
| Met Recurrence | DeepSeek-R1 | 4 | 17% | 54% | 28% |
| Met Recurrence | Gemini 2.5 Pro | 9 | 14% | 38% | 47% |
| Met Recurrence | GPT-4o | 4 | 22% | 58% | 29% |
| Met Recurrence | GPT-5 | 8 | 15% | 45% | 39% |

**Supplementary Table 4. RAG retrieval chunk rank by variable and LLM for dates of diagnosis and stage**

Distribution of retrieval rank for the chunk where the LLM found the cited evidence for extraction. Rank 1 means that this was the best match on the retrieval system (e.g. BM25 score). Tasks with earlier chunks such as the date of original diagnosis performed better than lower ranking chunks such as clinical stage.

| **Drug** | **Second oncologist** | **Gemini 2.5 Pro** | **GPT-5** | **GPT-4o** | **DeepSeek-R1** | **Research coordinator** |
| --- | --- | --- | --- | --- | --- | --- |
| **Missed Drugs** |  |  |  |  |  |  |
| Anastrozole | 1 | 1 | 2 | 3 | 2 | 31 |
| Paclitaxel | 5 | 6 | 6 | 5 | 5 | 10 |
| Letrozole | 0 | 0 | 0 | 0 | 3 | 21 |
| Exemestane | 0 | 0 | 0 | 1 | 1 | 22 |
| Cyclophosphamide | 0 | 5 | 4 | 4 | 7 | 7 |
| Doxorubicin | 1 | 4 | 4 | 3 | 5 | 7 |
| Docetaxel | 4 | 1 | 1 | 2 | 4 | 6 |
| Capecitabine | 0 | 0 | 0 | 1 | 10 | 6 |
| Talazoparib | 3 | 3 | 3 | 3 | 3 | 2 |
| Tamoxifen | 0 | 0 | 2 | 5 | 3 | 3 |
| Trastuzumab Deruxtecan | 1 | 2 | 2 | 2 | 3 | 3 |
| Clinical Trial | 1 | 1 | 1 | 1 | 5 | 3 |
| Eribulin | 0 | 0 | 0 | 0 | 6 | 3 |
| Atezolizumab | 1 | 1 | 1 | 1 | 3 | 1 |
| Palbociclib | 0 | 0 | 0 | 0 | 3 | 5 |
| Fulvestrant | 0 | 1 | 0 | 0 | 3 | 3 |
| Gemcitabine | 0 | 0 | 0 | 0 | 4 | 3 |
| Pembrolizumab | 1 | 1 | 1 | 1 | 3 | 0 |
| Carboplatin | 1 | 0 | 0 | 0 | 1 | 4 |
| Everolimus | 0 | 0 | 0 | 0 | 3 | 3 |
| **Added Drugs** |  |  |  |  |  |  |
| Aromatase Inhibitor | 0 | 0 | 0 | 1 | 0 | 51 |
| Docetaxel | 5 | 5 | 5 | 5 | 5 | 5 |
| Clinical Trial | 2 | 7 | 5 | 4 | 8 | 3 |
| Liposomal Doxorubicin | 4 | 4 | 4 | 4 | 4 | 5 |
| Exemestane | 2 | 6 | 6 | 4 | 4 | 0 |
| Paclitaxel | 5 | 2 | 2 | 4 | 2 | 5 |
| Nab-Paclitaxel | 0 | 5 | 4 | 4 | 4 | 1 |
| Fulvestrant | 0 | 4 | 4 | 4 | 3 | 2 |
| Capecitabine | 3 | 3 | 3 | 2 | 3 | 1 |
| Letrozole | 0 | 4 | 4 | 4 | 3 | 1 |
| Tamoxifen | 2 | 3 | 3 | 3 | 3 | 1 |
| Anastrozole | 1 | 4 | 3 | 3 | 3 | 0 |
| Gemcitabine | 1 | 2 | 3 | 4 | 1 | 3 |
| Atezolizumab | 0 | 2 | 2 | 2 | 1 | 6 |
| Eribulin | 0 | 3 | 3 | 4 | 1 | 1 |
| Carboplatin | 0 | 2 | 2 | 2 | 2 | 3 |
| Doxorubicin | 1 | 2 | 1 | 4 | 1 | 0 |
| Everolimus | 0 | 2 | 2 | 1 | 2 | 1 |
| Alpelisib | 0 | 2 | 2 | 1 | 2 | 0 |
| Cyclophosphamide | 1 | 1 | 1 | 3 | 1 | 0 |

**Supplementary Table 5. Most common missed and extra anti-cancer drugs across abstraction approaches.**
Most frequently missed and extra anti-cancer drugs relative to the first oncologist.

| **Line-level error metric** | **First oncologist** | **Second oncologist** | **Gemini 2.5 Pro** | **GPT-5** | **GPT-4o** | **DeepSeek-R1** | **Research coordinator** |
| --- | --- | --- | --- | --- | --- | --- | --- |
| Total lines | 716* | 704 | 740 | 714 | 725 | 668 | 689* |
| Temporally matched lines (start ±90 d) | - | 668 | 662 | 632 | 625 | 575 | 592 |
| Reference lines without a temporal match, per patient | - | 0.5 | 0.5 | 0.8 | 0.9 | 1.4 | 0.9 |
| Predicted lines without a temporal match, per patient | - | 0.4 | 0.8 | 0.8 | 1.0 | 0.9 | 1.0 |
| Patients with matching line count | - | 68 | 46 | 52 | 48 | 37 | 44 |

**Supplementary Table 6. Direction of error in systemic therapy abstraction relative to the expert oncologist reference.**
The table summarizes line-level error across the second oncologist, research coordinators, and all evaluated LLMs, including correctly matched lines, missed lines, and missed/extra lines relative to the expert oncologist reference. Lines are matched one-to-one by treatment start date within 90 days and do not require identical regimen composition; exact-regimen agreement (precision, recall, and F1) is reported in Table 2 and Supplementary Table 1. The * denotes that the research coordinators were compared against the oncologist reference censored on June 1, 2022 (681 of 716 lines); the 35 lines that began after that date were excluded.

| **Comparator** | **Jaccard, audited sample** | **Jaccard, overall cohort** | **F1, audited sample** | **F1, overall cohort** |
| --- | --- | --- | --- | --- |
| Second oncologist | 0.86 | 0.80 | 91% | 87% |
| Gemini 2.5 Pro | 0.77 | 0.70 | 85% | 80% |
| GPT-5 | 0.74 | 0.70 | 83% | 80% |
| GPT-4o | 0.80 | 0.69 | 87% | 79% |
| DeepSeek-R1 | 0.65 | 0.62 | 75% | 73% |

**Supplementary Table 7. Comparison of the Jaccard and F1 scores for the random sample of 20 patients reviewed for audit of the lines of therapy and the overall population**

| **Comparator** | **Main qualitative error patterns** | **Representative examples** |
| --- | --- | --- |
| Oncologist vs Oncologist | Disagreements were typically due to differences in line-grouping or omission of brief treatment courses by one of the oncologists. | One oncologist classified chemo-immunotherapy followed by maintenance immunotherapy as a single line, whereas the other classified these as two lines. In another case, one oncologist omitted an approximately 1-month course of endocrine therapy that was discontinued because of adverse effects. |
| Gemini 2.5 Pro | Similar to the inter-oncologist review, most differences involved line merging or grouping conventions. Brief treatment courses were missed less often than by the oncologists. Additional errors included inclusion of prescribed but apparently never initiated therapy. | In one case, an adjuvant chemotherapy switch due to an adverse event was called a single line of therapy even though it isn’t a standard regimen (both oncologists listed as two lines). In a separate case, Gemini collapsed serially trialed aromatase inhibitors that occurred over a short period of time into a single line.  It also caught very short courses of drugs that the oncologists often missed. For example, a ~1-month tamoxifen course was captured. Another error was adding an aromatase inhibitor that was prescribed, but later notes stated was never taken, as a line of therapy. In contrast, the oncologists and GPT-5 omitted this |
| GPT-5 | Errors included omission of a treatment course, differences in line splitting and merging, and interpretation of whether radiosensitizing capecitabine is a distinct line of therapy. | In one patient, GPT-5 omitted the entire adjuvant chemotherapy and endocrine therapy course, which was captured by all other models. Other differences were similar to the oncologists and Gemini 2.5 Pro: differences in grouping and splitting of drugs. An example of a split was GPT-5 counted a single dose of platinum as a separate line even though the treatment plan was to add a taxane and both oncologists considered this a single line. GPT-5 was also the only comparator to classify radiosensitizing capecitabine as a distinct line. Because the prompt did not explicitly specify how radiosensitizing therapy should be handled, this was considered primarily an interpretation difference rather than a clear abstraction error. |
| GPT-4o | In addition to line-splitting and merging differences, errors included missing or incomplete dates, internally inconsistent dates, incorrect treatment ordering, and missed therapies. Overall abstraction quality was qualitatively lower than for Gemini 2.5 Pro and GPT-5 despite identification of most anti-cancer drugs. | GPT-4o more frequently left dates incomplete, for example assigning only a month or year rather than an exact date. In one patient, it recorded a stop date that preceded the start date, shifting the apparent position of the line and disrupting the treatment sequence, although the drugs were correct. The post-abstraction pipeline did check chronology across lines but did not test whether a stop date preceded the start date within the same line. GPT-4o was also the only model to miss adjuvant tamoxifen in one patient. In another case, it split aromatase inhibitor plus abemaciclib into two sequential therapies, whereas all other comparators treated the combination as a single line. |
| DeepSeek-R1 | DeepSeek had unique errors including an insertion of a drug and return of an empty JSON packet (presumably due to truncation in the Stanford SecureGPT API) in patients with complex treatment histories. There were also errors in chronology, despite extraction of correct dates, as well as standardization errors, and the expected line splitting or merging. | The most consequential error was truncation in a reviewed patient with > 10 lines of therapy where Deepseek returned no metastatic therapies. Despite retry attempts x3 as part of the pipeline for empty JSON and increasing token allowances with each retry that included the model max, they still returned empty. We suspect this was due to limitations on the institution’s API. DeepSeek-R1 also occasionally placed treatment lines out of chronological order despite extracting the underlying dates correctly--essentially garbling the standardized output it was asked to produce even when it retrieved the correct evidence. In one reviewed case, it added a drug that was not present in the chart: the supporting quotation identified an aromatase inhibitor and palbociclib, but the standardized output included these two drugs and abemaciclib. This is not a regimen but given the semantic similarity of two adjacent CDK4/6 inhibitors suggests that EOS token was not selected when it should have been. It also classified a single dose of an ovarian-suppression agent as a treatment line despite explicit instructions not to treat ovarian suppression as a separate line. |
| Overall Qualitative Summary | For the oncologists, GPT-5, and Gemini 2.5 Pro, most discrepancies reflected judgment calls: how to split or merge therapies into individual lines. For example, if TCHP followed by HP maintenance was one or two lines of therapy. These two LLMs often captured short courses of agents (radiosensitization, < 1 month of endocrine therapy) that an oncologist would miss but did also miss therapies as well.  GPT-4o and DeepSeek R1 had these failures and an additional set of qualitatively different failure modes. While differences in line splits and merges were seen, we saw response truncation, unsupported drug insertion, internally inconsistent dates, and chronology errors. Many of these errors may be addressable through additional validation rules and guardrails. However, the qualitative difference suggests that general model improvement may reduce these errors. | |

**Supplementary Table 8. Qualitative review of differences in the classification of lines of therapy**

| **Outcome** | **Covariate** | **Expert-derived HR (95% CI)** | **Gemini 2.5 Pro-derived HR (95% CI)** | **GPT-5-derived HR (95% CI)** | **Cochran Q P value** |
| --- | --- | --- | --- | --- | --- |
| Overall survival | Stage 4 vs stages 1-3 | 2.67  [1.26 - 5.63] | 2.58  [1.25 - 5.32] | 2.85  [1.38 - 5.86] | p = 0.95  p = 0.90 |
| Overall survival | Hormone receptor-negative vs hormone receptor-positive | 2.06  [1.14 - 3.73] | 1.60  [0.87 - 2.91] | 1.66  [0.91 - 3.00] | p = 0.55  p = 0.61 |
| Recurrence-free survival | Stage 3 vs stages 1-2 | 2.34  [1.45 - 3.77] | 2.18  [1.33 - 3.55] | 2.19  [1.35 - 3.56] | p = 0.84  p = 0.85 |
| Recurrence-free survival | Hormone receptor-negative vs hormone receptor-positive | 2.07  [1.31 - 3.29] | 1.61  [1.01 - 2.56] | 1.79  [1.13 - 2.83] | p = 0.45  p = 0.66 |

**Supplementary Table 9. Cox proportional hazards model estimates using expert-derived and LLM-derived data.**
Univariate hazard ratio estimates and 95% confidence intervals for survival estimates using expert vs LLM datasets. In the Cochran Q column, the top P value is for Gemini 2.5 Pro and the lower one for GPT-5.

| **Variable** | **Definition** |
| --- | --- |
| **Clinical variable definitions** | |
| Clinical stage | When an explicit final stage group was documented by the clinician, this was used. However, if not documented it was inferred from the available data for LLMs and human reviewers. We only examined the primary clinical stage (1, 2, 3, or 4) and not substage. We used the AJCC 7th edition, but did include pathologic upstaging when postoperative pathology increased the initial documented stage and there was no neoadjuvant therapy delivered. This staging convention was applied consistently across expert abstraction and LLM prompts. |
| Recurrence | Locoregional recurrence was defined as recurrence in the ipsilateral breast, chest wall, axillary lymph nodes, supraclavicular lymph nodes, or internal mammary lymph nodes after a prior breast cancer diagnosis and completion of initial definitive neoadjuvant and locoregional therapies with concurrent negative imaging for distant disease. Distant recurrence was defined as recurrence at any other site. When assigning recurrence dates, pathology was prioritized when available, but clinical context was used as well. |
| Multiple primary breast cancers | For patients with multiple primary breast cancers, abstraction focused on the malignancy that was clinically dominant at recurrence or metastatic progression. If a patient had synchronous bilateral primaries with differing receptor profiles but later developed biopsy-proven metastatic disease matching one subtype, abstraction was anchored to the cancer that appeared to have become metastatic. If a patient had multiple distinct invasive primaries without recurrence, the later event was abstracted if it was judged to represent a distinct primary rather than locoregional recurrence (different subtype, contralateral breast, etc.). Ductal carcinoma in situ was not considered an invasive recurrence event. |
| **Tumor subtype and stage** | |
| Hormone receptor and HER2 definitions | Hormone receptor status was derived from estrogen receptor and progesterone receptor results. Priority was given to pathology, with a fallback to clinical documentation when original pathology was unavailable in the chart. Tumors were classified as hormone receptor-positive if the original core biopsy or surgical specimen documented estrogen receptor or progesterone receptor staining greater than 10%, or an equivalent clinical interpretation of positivity (i.e. “positive” written in the chart or treatment on HR-positive algorithms). Tumors were classified as hormone receptor-negative if both markers were negative.  HER2 status was adjudicated using CAP guidelines when pathology was available. If the original pathology was unavailable, the clinical interpretation and treatment decisions were used to assign HER2 status. |
| Date of death | Death date was assigned only when explicitly documented. For example, if a patient was discharged to hospice in 2016 with no further notes in the chart, and no documented date of death then this variable was left blank. For this study, we did not have merged death records from the state or registry. |
| **Systemic therapy** | |
| Line definitions | Lines of therapy were defined according to oncologist adjudication rather than algorithmic inference. Maintenance transitions and sequential regimens were documented using a consistent convention. For example, AC-T was recorded as a single line as “doxorubicin + cyclophosphamide-->paclitaxel”, and TCHP followed by maintenance trastuzumab plus pertuzumab was recorded as “TCHP >-> HP” with --> for transitions in a single line to the second portion of the therapy and >-> to denote maintenance transitions. For combination therapies we looked at starts within 90 days. For example, if letrozole + palbociclib were used together even if not started on the identical day these were inferred as a singular line of therapy with the start date anchored to the first drug started. A therapy had to be taken to count as a line; drugs that were prescribed or planned but never started were not lines, and listing a never-initiated drug as a line was therefore treated as an abstraction error. |
| Clinical trials | For clinical trials, the abstracted record included the drugs used when known. However, because randomization, blinding, and evolving compound names could complicate exact line assignment, the line itself was recorded as “clinical trial” and we did not look at the individual drugs for these lines. |
| Drug name normalization and class collapsing | The abstraction target for the drug- and line-level analyses was the set of regimen-defining anti-cancer agents that were initiated—cytotoxic chemotherapy, endocrine therapy, HER2-directed and other targeted agents, and immunotherapy. Supportive-care medications (ovarian suppression such as goserelin, bone-modifying agents such as denosumab and bisphosphonates, growth factors, and antiemetics) were not part of this target and were removed before analysis. For both line-based and drug-based analyses, we used a standardized normalization dictionary to harmonize drug names across abstraction sources. Prompts provided instructions to the LLMs on how to normalize outputs. There was an additional cleaning script used for all the outputs (human and LLM) run prior to analysis that corrected human typos, stripped out supportive medications including ovarian suppression (e.g., goserelin) and bone-modifying agents (e.g., denosumab) and normalized clinical trial lines to ‘clinical trial’.  Because research coordinators often wrote “aromatase inhibitor” and other times wrote the drug name, we did an additional post-hoc sensitivity analysis where we collapsed all aromatase inhibitors (anastrozole, letrozole, exemestane) into a single aromatase inhibitor category and individual taxanes (paclitaxel, docetaxel, and nab-paclitaxel) into a single taxane category. |
| Treatment intent | Treatment intent was abstracted as curative vs palliative with the major distinction being derived for pre-distant metastatic disease vs post-metastatic disease diagnosis. Locoregional therapy was considered curative in intent. |
| Reason for discontinuation if a line of therapy | Reason for discontinuation were categorized into the following for clinicians and the LLM: disease progression, completed planned neoadjuvant course, completed planned adjuvant course, toxicity, patient preference, clinician choice, trial protocol ended, hospice enrollment, patient death, ongoing (only for active therapy at the time of censor in January 2026). For the primary analysis, these categories were collapsed into progression-related discontinuation (disease progression, hospice enrollment, or death) vs other reasons, which included planned treatment completion, toxicity, patient preference, clinician choice, and trial completion. |
| **Genetic testing** | |
| Reference standard | For germline BRCA1/2 mutation status and somatic ESR1 and PIK3CA mutation status, linked genetic testing records within the institutional database served as the reference standard rather than clinician abstraction (i.e. the PDFs directly from the companies). The primary genetics analysis evaluated detection of a pathogenic variant when present in the linked testing record. For this binary mutation-presence analysis, cases without evidence of the specified mutation were categorized as mutation not identified, regardless of whether testing was negative or the specific alteration had not been assessed. |

**Supplementary Table 10. Clinical variable definitions used for expert abstraction and LLM prompting.**

| **Clinical variable** | **Abstraction method** |
| --- | --- |
| Date of initial breast cancer diagnosis | Identified from pathology reports and clinical notes using a task-specific retrieval and LLM extraction workflow designed to prioritize the original invasive breast cancer diagnosis and pathology provenance and enforce consistency with later recurrence dates. |
| Date of birth | Identified primarily from demographic records, with broader chart review used only when demographic data were incomplete or unavailable. |
| Date of death | Identified primarily from demographic records when available and otherwise from clinical documentation near the end of the chart. |
| Date of first locoregional recurrence | Extracted from longitudinal clinical documentation using a date-focused abstraction workflow, with consistency checks to ensure that recurrence occurred after the initial diagnosis. |
| Date of first distant metastatic diagnosis | Extracted from pathology reports and clinical notes using a date-focused abstraction workflow designed to identify the first documented distant metastatic event and preserve temporal consistency with the original diagnosis. |
| Clinical stage at diagnosis | Determined from pathology reports and peri-diagnostic clinical documentation using a staged extraction workflow focused on the original cancer presentation. |
| Hormone receptor and HER2 status | Derived from diagnostic pathology when available and otherwise from clinical documentation anchored to the original diagnosis. |
| Primary treating oncologist | Determined from structured visit information and clinical documentation to identify the physician responsible for most medical oncology care for a given patient in the outpatient setting. |
| Germline BRCA1/2 mutation status | Derived from clinical documentation of germline testing and binarized as pathogenic or likely pathogenic BRCA1/2 variant present vs other. Variants of uncertain significance were classified as negative. |
| Somatic PIK3CA and ESR1 mutation status | Derived from clinical documentation of somatic testing and binarized as mutated vs other. |
| Lines of systemic anti-cancer therapy | Reconstructed from clinical notes and medication records using a longitudinal abstraction workflow designed to identify regimen composition, treatment timing, intent, and reason for discontinuation across curative-intent and recurrent or metastatic settings. |

**Supplementary Table 11. Overview of methods for Variable Abstraction**

High-level summary of the abstraction approach used for each clinical variable. Each task used a variable-specific retrieval and LLM extraction workflow tailored to the expected location and structure of the relevant information in the chart. Detailed implementation and prompt logic are available in the code repository.
